# Performance evaluation and benchmarking across 16 large language model configurations on a large real-world emergency department triage data set

**DOI:** 10.64898/2026.05.28.26353935

**Authors:** Leo Benning, Anja Hirsch, Matthias Gröschel, Tobias Röschl, Martin Spott, Felix Patricius Hans, Tim Urban, Hans-Jörg Busch, Alexander Meyer, Julian Madrid

## Abstract

**Background:** Emergency department (ED) triage is a decision process that determines patient prioritization and resource allocation. Large language models (LLMs) have been proposed as decision-support tools for triage, yet most evaluations rely on curated datasets. Evidence from real-world clinical environments remains limited. The objective of this project was to evaluate the performance and reproducibility of multiple contemporary large language models for Emergency Severity Index (ESI) classification and sectoral allocation (ED vs. urgent care practice, UCP) using a real-world triage dataset.

**Material and Methods:** Retrospective cross-sectional benchmarking study conducted at a tertiary academic ED in Germany with an integrated central point of assessment (CPA). The study included all 16,107 walk-in encounters between October 18, 2023, and February 27, 2024. Sixteen configurations were compared on the first 2,000 encounters; GPT 5.1 with high reasoning effort was additionally evaluated on all 16,107 encounters. Data were collected from a structured clinical decision support system capturing presenting complaints, vital signs, and triage decisions recorded by specialized nursing staff.

**Results:** The primary outcome was the agreement between LLM-predicted and nurse-assigned ESI levels measured using quadratic-weighted Cohen’s κ. Secondary outcomes included sectoral assignment agreement, misclassification patterns (over- and under-triage), confidence–agreement comparisons, and output reproducibility. In the primary analysis of all 16,107 encounters, GPT 5.1 with high reasoning effort achieved quadratic-weighted κ = 0.5031 (95% CI 0.4904–0.5158). In the exploratory comparison of 16 configurations on the shared 2,000-encounter subset, quadratic-weighted κ ranged from 0.180 to 0.860. The Stepwise approach, combining structured model assessments with deterministic ESI mapping, achieved κ_qw = 0.860 (95% CI 0.841–0.878) and 82.70% exact agreement. Most models demonstrated moderate or lower agreement. Sectoral assignment agreement (i.e. ED vs. urgent care practice, UCP) was uniformly low (κ ≤ 0.304). Reproducibility testing revealed substantial variability in 23% of cases, indicating non-deterministic output behavior for clinically relevant decisions.

**Conclusions:** Current large language models demonstrate heterogeneous and generally limited performance in real-world emergency triage tasks. The Stepwise approach achieved the highest agreement. Before clinical implementation, confidence estimates, reliability, and workflow integration require prospective validation in regulatory-compliant clinical settings.

## Introduction

Emergency departments (EDs) are critical entry points to medical care in most healthcare systems. Equipped to assess severity of presentations, provide critical acute care where needed and discharge patients to either inpatient care, outpatient specialist services, or primary care, emergency care delivery in EDs is characterized by high numbers of patient contacts with often unfiltered presenting complaints and little time to decide which patient requires what type of care. Simultaneously, however, the identification of the appropriate type of care is critical to both individual patients and the ED itself, as ill-allocated resources have immediate effects on patient outcomes ^1^ and the phenomena around ED crowding ^2,3^. This challenge has become more pronounced over the past years, as EDs have seen increasing patient numbers, exemplarily in Germany and the United States ^4–6^.

### Triage systems

The fundamental features of emergency medicine (EM) have led to the development of triage algorithms supporting the initial assessment of patients regarding their chief complaints, vital signs and indications of an immediate threat to life. Although subject to a long and ongoing discussion, triage is typically performed by specialized nursing staff ^7–9^. Conventional triage systems are most often used within EDs and help to assess acuity prior to treatment. Most established are the Manchester Triage System (MTS) and the Emergency Severity Index (ESI) ^10,11^, but also the Australasian Triage Scale (ATS), the Taiwan Triage System (TTS) and the Canadian Triage and Acuity Scale (CTAS) have received attention in comparative studies on the effectiveness of the different approaches ^12,13^. As employed in the center assessed in this study, we will focus on ESI here, while extensive literature on triage systems has been published elsewhere ^14,15^.

ESI is based on a stepwise assessment ^16^: First, the need for any acute and life-saving interventions is assessed. If any are required, the highest triage category (i.e. ESI 1) is assigned. If none are required, but a high-risk situation is anticipated, ESI 2 is assigned. If both are not given, the number of anticipated resources is assessed. Although ESI is well established, concerns about inter-rater reliability and the risk of under-triage (i.e. assignment of inappropriately low triage categories) have long been discussed ^17,18^. More recently, the effects of ED crowding (i.e. episodes during which the need for emergency medical care exceeds available resources in the ED, the hospital, or both ^19^) and nursing staff shifts on triage have been explored: During crowding, process failures (i.e. no triage categories assigned) and triage delays occurred significantly more frequently than during non-crowding episodes ^20^. Similarly, nursing staff appeared to become more lenient in their assessment of the acuity of presentations as their shift progresses ^21^, potentially due to highly repetitive tasks and effects of fatigue. These findings around a context-sensitive and operator-dependent performance variability of triage assessment are important shortcomings of the current practice of triage that warrant further consideration.

### Central point of assessment

To better coordinate patient influx in EDs and to allocate available healthcare resources more effectively, recent healthcare policy efforts in Germany focused on reorganizing acute care delivery ^22^. A core principle of this reform is a structured patient guidance by steering patients to the most appropriate level of care before they self-admit themselves to an emergency department. For patients presenting in person, integrated emergency care centers (IEC) provide a central point of assessment (CPA) as a single entry point upstream of the combined ED and ambulatory urgent care practice (UCP). At this CPA, a standardized assessment assigns patients to the appropriate sector (i.e. UCP or ED), thereby moving triage to an earlier point in the patient journey and enabling diversion of low-acuity patients to less resource-intensive care pathways ^23–25^.

At our center, a web-based clinical decision support system (CDSS) used at the CPA to assign acuity (ESI levels) and appropriate care sector is operated by specialized triage nursing staff and captures the patient’s chief complaint according to the Canadian Emergency Department Information System Presenting Complaint List (CEDIS-PCL) ^25,26^, along with vital signs and structured items to assess the patient’s acuity and the appropriate care sector. Based on these inputs, the CDSS generates a suggestion for i) Emergency Severity Index (ESI) and (ii) for sectoral allocation within the IEC (i.e. UCP or ED). Yet, these suggestions only provide cues for decision support while the definitive triage and allocation decision remains with the triage nursing staff.

### LLM-supported triage and allocation decision support

To address the shortcomings of context-sensitive and operator-dependent triage performance in the context of our IEC, our objective was to assess the performance of a broad set of state-of-the art Large Language Models (LLMs) on a real-world data set recorded at our center’s CPA to apply i) the ESI rules to provide a recommendation for the ESI category of the respective patient and ii) suggest the appropriate sectoral assignment (i.e. UCP or ED) against the recorded assessment by specialized triage nursing staff. Previous LLM triage evaluations have included fictional case vignettes ^27,28^, case series of real emergency-department cases ^29^, and curated triage data sets ^30,31^ like MIMIC-IV ^32^ or NEJM Healer ^33^. Before this background, we evaluate the real-world applicability of LLMs in an acute-care setting and highlight practical challenges related to implementation, validation, and the regulatory context.

## Methods

### Data collection

This work presents a retrospective cross-sectional study conducted at a tertiary level academic medical center. The study retained all 16,107 walk-in encounters, including pediatric age-band records, with recorded dates from October 18, 2023, through February 27, 2024. Emergency Medical Services (EMS) arrivals followed a separate admission pathway and were not included in this analysis. All data for this study were obtained from the custom-built CDSS that is operated at the CPA of our medical center ^25^. It resembles an Electronic Health Record (EHR) for patient records before administrative registration. All variables used are listed in Annex A. In brief, at the CPA patients draw a queuing ticket upon entering the medical center and are called up by the specialized nursing staff at the CPA. After arrival of the patients in the triage room, staff assesses the patients’ chief complaint based on CEDIS-PCL ^26^, collects vital signs according to the ABCDE-standard ^34^. The nursing staff is free to add additional free-text comments, structured risk factors or injuries if deemed necessary information for the subsequent care provider. All data is entered in German. Based on the structured data input, the CDSS provides a rule-based suggestion for both ESI category and sectoral assignment (i.e. UCP or ED), which the nursing staff can override in case they deem the suggestion inappropriate or unavailable. The proposal of the CDSS together with the final decision by the nursing staff is included in a triage report card that can either be printed, or accessed via the personalized QR code to be available in the subsequent care sector. For this study, only the final decision by the nursing staff is used as the reference. The patient flow is illustrated in Figure 1. Annex B describes the triage report card.

**Figure 1:**
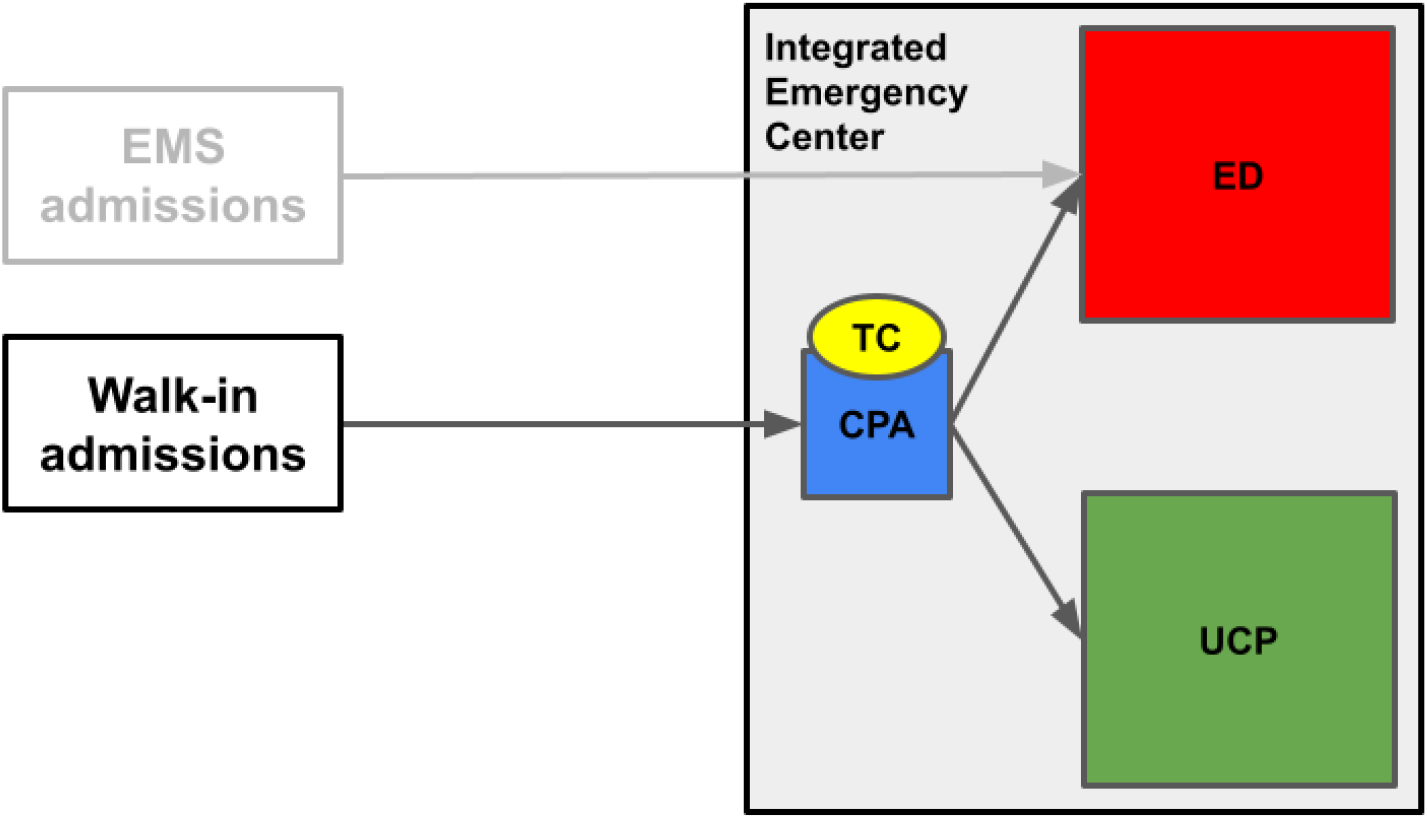
Illustration of the patient flow. Unscheduled walk-in admissions to the Integrated Emergency Center (IEC, dark grey) go through the Central Point of Assessment (CPA, blue), where ESI level and care sector (i.e. UCP or ED) are assigned by specialized nursing staff using the CDSS TriageClient (TC, yellow) for documentation and data storage. This step is completed prior to the administrative admission of the patient. Upon assessment, patients receive a triage report card (Annex B). They are either referred to the Emergency Department (ED, red) or the Urgent Care Practice (UCP, green), where the administrative admission and subsequent treatment takes place. Emergency Medical Services admissions (EMS, light grey) are not subject of this study but are displayed in this illustration for context.

### Analytical Methods

#### LLM evaluation protocol

Fourteen individual model configurations, one ensemble strategy and a Stepwise approach were evaluated (Annex C). Models were selected to represent a broad cross-section of contemporary LLM families and reasoning configurations. The common input for individual configurations comprised available recorded clinical variables in a structured format and excluded the nurse-assigned ESI and sector (Annex A). Models were instructed to return an ESI classification (i.e. ESI 1–5), a sectoral assignment (i.e. UCP or ED), and a self-reported verbalized confidence level (0–100%). The standard prompt included ESI rules and three demonstrations; the Council and Stepwise workflows received workflow-specific prompts. Available prompt templates and configuration labels are reported in Annexes C and D. The Stepwise prompt instructed the model to return a negative danger-zone flag when the available information was insufficient to assess the relevant thresholds.

For the primary analysis, GPT 5.1 with high reasoning effort was prompted on the full cohort of 16,107 encounters to assess large-sample performance. For exploratory analyses, each model assessed (Annex C) independently classified the first 2,000 cases to enable a comparative assessment of the models. Outcomes were compared to final triage decisions as assigned by specialized triage nursing staff and recorded in the CDSS.

Two additional strategies were evaluated. The “Council of Models” approach^35^ aggregated predictions from nine individual models (GPT 5.2, DeepSeek V3.2, Kimi K2, Llama 4 Maverick, Mistral Large 3, GPT 5 Pro, Claude Opus 4.5, Gemini 3 Pro, and Grok 4) for each patient, with the final ESI-level determined by GPT 5.2 as the chairman model through review and synthesis. The Stepwise approach used GPT 5.1 with high reasoning effort to generate structured intermediate assessments corresponding to the decision points of the ESI algorithm. These assessments were converted into a final ESI classification using a deterministic mapping of the ESI-based decision rules. The Stepwise prompts and deterministic decision rules are provided in Annex D^16^.

All models were accessed via application programming interfaces (APIs); no local model training or fine-tuning was performed.

#### Reproducibility experiment

To quantify the stochastic variability of LLM outputs, GPT 5.1 (high reasoning effort) was queried 100 times with identical input for each of the first 100 encounters, yielding 10,000 classifications. The experiment used repeated calls for the same records.

#### Statistical analysis

Analyses of the archived outputs used Python 3.12.14, NumPy 2.3.5 and pandas 2.2.3, with agreement estimates independently checked using scikit-learn 1.8.0. The primary outcome was the agreement between each LLM-predicted ESI level and the nurse-assigned ESI level at the CPA. Because ESI is an ordinal five-level scale, we computed Cohen’s κ with three weighting schemes: unweighted κ, linear-weighted κ (κ_lin), and quadratic-weighted κ (κ_qw) ^36,37^. Quadratic-weighted κ was selected as the primary measure because it penalizes larger disagreements disproportionately, and because it is related to the intraclass correlation coefficient ^37,38^. Agreement was interpreted according to the Landis and Koch scale: <0.00 poor, 0.00–0.20 slight, 0.21–0.40 fair, 0.41–0.60 moderate, 0.61–0.80 substantial, and 0.81–1.00 almost perfect ^39^. 95% confidence intervals (CI) were estimated using a nonparametric percentile bootstrap with 10,000 encounter-level resamples and a fixed random seed of 42 ^40^. Agreement within ±1 ESI level was defined as the proportion of paired cases in which the absolute difference between the model-predicted and nurse-assigned ESI category was ≤ 1. Agreement patterns were additionally visualised using row-normalized confusion matrices and error-distance distributions, where error distance was defined as the absolute difference between model-predicted and nurse-assigned ESI level.

The signed deviation (LLM-ESI level minus nurse-ESI level) was computed for each encounter. Negative deviations indicate over-triage, positive deviations indicate under-triage. We report the proportions of encounters over-triaged and under-triaged and the mean signed deviation. These terms denote disagreement with recorded nurse ESI. High-acuity misses were nurse ESI 1–2 encounters assigned model ESI 3–5.

Sectoral assignment agreement was assessed using unweighted Cohen’s κ and proportions of misclassification to either ED or UCP conditional on the nurse-assigned ED or UCP destination. We assessed verbalized confidence, defined as the model’s self-reported certainty ^41^. The prompt requested generic certainty given data completeness and ambiguity alongside the ESI and destination decisions; it did not request the probability of exact agreement with nurse-assigned ESI. Confidence analyses therefore describe associations and numerical differences between this certainty score and exact ESI agreement. Confidence scores were expressed on a 0–1 scale. The confidence–agreement gap was calculated as mean confidence minus the proportion of exact ESI matches. We also divided confidence into ten equal-width bins. The weighted absolute bin-level gap was calculated as the sum of absolute differences between mean confidence and exact agreement within each bin, weighted by the proportion of encounters in that bin. This calculation follows the mathematical formula for expected calibration error ^42^ but is interpreted here as a descriptive discrepancy rather than a measure of probability calibration. Confidence accuracy (CA) was calculated using our previously introduced metric ^43^ as mean confidence among exact ESI matches minus mean confidence among ESI mismatches.

For the reproducibility experiment, we computed per-encounter modal ESI frequency and number of unique ESI levels. Multi-rater reliability was summarised using Fleiss’ κ ^44^. Modal aggregation across all 100 runs was compared with mean single-run exact agreement; tied modes were assigned the smaller ESI number. Access type, reasoning labels and public model metadata ^45^are reported descriptively (Annex C).

All confidence intervals are reported at the 95 % level. No adjustments for multiple comparisons were applied, as the analyses were intended as estimation rather than hypothesis testing ^46^.

Analyses included encounters with usable model outputs for the respective endpoint. Encounters with missing outputs were excluded from the affected endpoint analysis but retained in other analyses when the required outputs were available. Endpoint-specific analysis denominators are reported in the corresponding tables.

### Reporting, Open Science and Ethical Approval

The reporting of this article follows the TRIPOD-LLM guideline ^47^ and the respective checklist is available in Annex E. The study was approved by the ethics committee of the University of Freiburg on July 9, 2024, with reference number 24-1052-S1-retro. The requirement for individual informed consent was waived. Beyond individual funding, as outlined in the respective section below, this study was funded with internal resources. FPH declares a potential conflict of interest as a stakeholder in Blackpine Medical GmbH, the manufacturer of the CDSS used to record triage data. This relationship had no influence on the preparation of the manuscript. All other authors do not declare any conflicts of interest. All study documentation and analysis code may be requested from the authors, subject to institutional data protection requirements. There was no public involvement for this study.

## Results

### Study cohort and data availability

The analysis cohort comprised 16,107 unscheduled walk-in encounters recorded at a tertiary level academic medical center between October 18, 2023, and February 27, 2024. Each configuration was evaluated on the first 2,000 encounters, with one model (GPT 5.1, high reasoning effort) additionally evaluated on the full cohort. Grok 4 returned incomplete responses for some encounters. Usable outputs were available for ESI agreement analysis in 2,000 encounters, sectoral assignment analysis in 1,998 encounters, and confidence analysis in 1,997 encounters. The nurse-assigned ESI distribution reflected the following acuity distribution in the full cohort: ESI 1, 0.1 % (n = 20); ESI 2, 11.9 % (n = 1917); ESI 3, 36.8 % (n = 5928); ESI 4, 38.6 % (n = 6224); ESI 5, 12.5 % (n = 2018) (Table 1a). The majority of patients (64.1 %) were directed to the ED, with 35.9 % directed to the UCP (Table 1b). The distribution of chief complaints shows a high proportion of trauma and musculoskeletal complaints (7/20 of the most prevalent chief complaints accounting for 29.8% of all presentations) as well as a long-tail distribution with 138 chief complaints accounting for 39% of all presentations (Table 1c). Annex C summarises the characteristics of all evaluated configurations. The 2,000-record comparison sample is outlined in Annex F.

**Table 1a:**
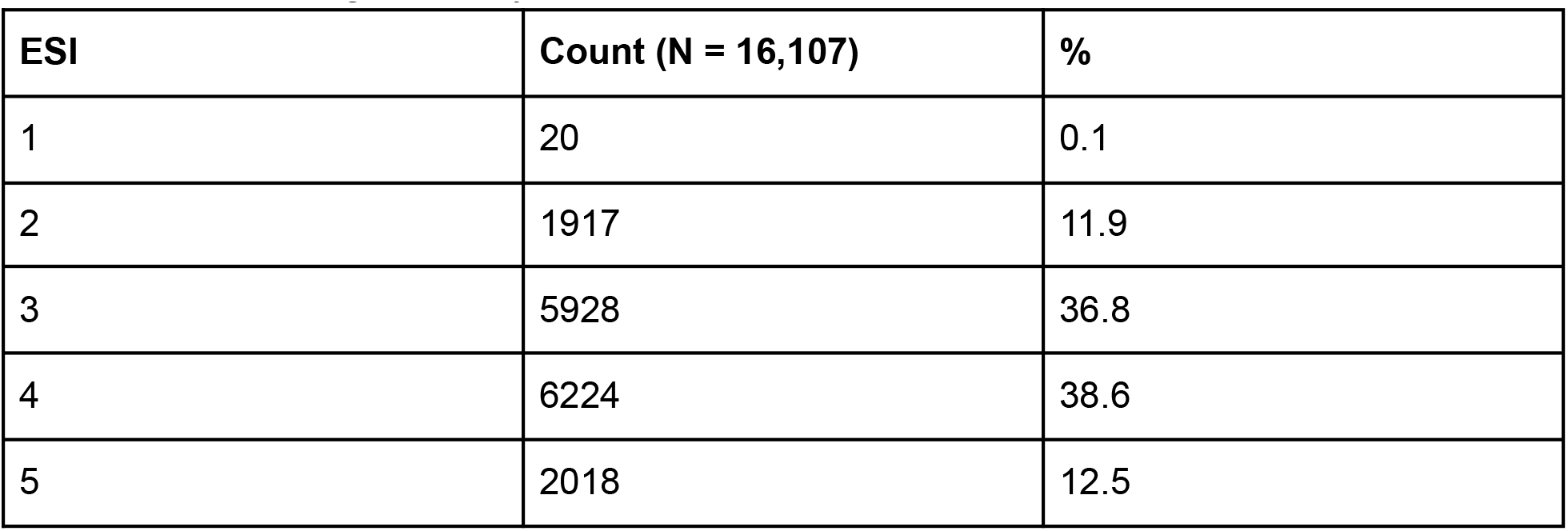
Nurse-assigned acuity distribution of encounters at the Central Point of Assessment.

**Table 1b:** Nurse-assigned sectoral allocation at the Central Point of Assessment.

| Sector | Count (N = 16,107) | % |
| --- | --- | --- |
| ED | 10321 | 64.1 |
| UCP | 5786 | 35.9 |

**Table 1c:** Distribution of the 20 most common chief complaints at the Central Point of Assessment.

| Chief Complaint | CEDIS code | Count (N = 16,107) | % |
| --- | --- | --- | --- |
| Lower extremity pain | 555 | 1308 | 8.1 |
| Upper extremity pain | 554 | 1049 | 6.5 |
| Abdominal pain | 251 | 1034 | 6.4 |
| Upper extremity injury | 556 | 607 | 3.8 |
| Laceration/Puncture | 704 | 576 | 3.6 |

| **Chief Complaint** | **CEDIS code** | **Count (N = 16,107)** | **%** |
| --- | --- | --- | --- |
| Backpain | 551 | 571 | 3.5 |
| Influenza-like illness | 661 | 567 | 3.5 |
| UTI complaints | 307 | 419 | 2.6 |
| Headache | 404 | 418 | 2.6 |
| Lower extremity injury | 557 | 414 | 2.6 |
| Dental/gum problem | 101 | 409 | 2.5 |
| Chest pain - cardiac features | 003 | 361 | 2.2 |
| Localized swelling/redness | 709 | 291 | 1.8 |
| Upper respiratory infection symptoms | 154 | 282 | 1.8 |
| Head injury | 407 | 277 | 1.7 |
| Vertigo | 403 | 273 | 1.7 |
| Chest pain - non-cardiac features | 004 | 251 | 1.6 |
| Fever | 852 | 245 | 1.5 |
| Prescription/medication request | 861 | 242 | 1.5 |
| Abnormal lab values | 863 | 234 | 1.5 |
| Other 138 complaints | n/a | 6279 | 39 |

The CDSS allows the collection of vital signs in the format of Airway (i.e. open or compromised), Breathing (e.g. breathing rate, peripheral oxygen saturation), Circulation (e.g. heart rate, blood pressure, recapillarization), Disability (e.g. signs of paralysis, impaired alertness) and Exposure (temperature or pain) according to the ABCDE framework ^34^ (Annex A). Data entries are not mandatory and blank categories are, hence, possible. As availability of appropriate data for each category are, however, crucial for the assessment of both acuity and appropriate care sector, the distribution of any data entry for each clinical domain is displayed in Table 2.

**Table 2:** Distribution of available clinical domains of the ABCDE framework^34^ in the recorded data (Annex A). Availability means at least one constituent field.

| Clinical Domain | Count available (of N = 16,107) | % available |
| --- | --- | --- |
| Airway | 8,420 | 52.3 |
| Breathing | 13,709 | 85.1 |
| Circulation | 14,025 | 87.1 |
| Disability | 1,056 | 6.6 |
| Exposure | 13,766 | 85.47 |

Respiratory rate was documented in 347/16,107 encounters (2.15%), total Glasgow Coma Scale in 1,036/16,107 (6.43%), glucose in 21/16,107 (0.13%) and pain score in 3,842 (23.85%). These frequencies describe documentation, not whether bedside measurements were performed.

### ESI classification agreement

Quadratic-weighted kappa values ranged from 0.180 (95 % CI 0.149–0.211; GPT 5 Nano, low reasoning) to 0.860 (95 % CI 0.841–0.878; Stepwise approach) across 16 model configurations (Table 3a). Only one strategy (the Stepwise approach) achieved agreement above κ_qw = 0.60. Seven configurations reached moderate agreement (κ_qw 0.41–0.60): the Council of Models ensemble (κ_qw = 0.5161, 95 % CI 0.4834–0.5477), GPT 5.1 with high reasoning effort on both the full cohort and the 2,000 encounter sub-sample (κ_qw = 0.5031, 95% CI 0.4904–0.5158 and κ_qw = 0.4692, 95% CI 0.4321-0.5056, respectively), GPT 5.1 with no reasoning (κ_qw = 0.4836, 95 % CI 0.4481-0.518), Claude Opus 4.5 (κ_qw = 0.4809, 95 % CI 0.4443-0.5163), Gemini 3 Pro (κ_qw = 0.4656, 95% CI 0.4292-0.5011), GPT 5 Pro (κ_qw = 0.435, 95% CI 0.396–0.472) and Mistral Large 3 (κ_qw = 0.418, 95% CI 0.380–0.455). Six configurations demonstrated fair agreement (κ_qw 0.21–0.40), and two (both GPT 5 Nano variants assessed) achieved only slight agreement (κ_qw < 0.20) (Table 3a). Within-±1-level agreement ranged from 62.25 % (GPT 5 Nano) to 97.85 % (Stepwise approach), indicating that the majority of disagreements were adjacent-level ESI misclassifications (Table 3b).

**Table 3a:** Agreement analysis of models assessed versus nurse-assigned acuity classifications based on ESI. 95% CIs use 10,000 encounter-level bootstrap resamples.

| **Model** | **N** | **$\kappa$ (quadratic)** | **95%-CI** |
| --- | --- | --- | --- |
| Stepwise approach<br>(GPT5.1 high reasoning) | 2000 | 0.8603 | 0.8415-0.8780 |
| Council of Models | 2000 | 0.5161 | 0.4834-0.5477 |
| GPT5.1 (no reasoning) | 2000 | 0.4836 | 0.4481-0.518 |
| Claude Opus 4.5 | 2000 | 0.4809 | 0.4443-0.5163 |
| GPT5.1 (high reasoning) | 2000 | 0.4692 | 0.4321-0.5056 |
| Gemini 3 Pro preview | 2000 | 0.4656 | 0.4292-0.5011 |
| GPT5 Pro (high reasoning) | 2000 | 0.4348 | 0.3955-0.4725 |
| Mistral Large 3 | 2000 | 0.4182 | 0.3801-0.4545 |
| GPT5.2 (xhigh reasoning) | 2000 | 0.3985 | 0.3606-0.4358 |
| Kimi K2 (thinking) | 2000 | 0.361 | 0.327-0.3947 |
| DeepSeek V3.2 | 2000 | 0.3607 | 0.3236-0.3974 |
| Llama 4 Maverick | 2000 | 0.3307 | 0.2908-0.3694 |
| Grok 4 | 2000 | 0.3223 | 0.2869-0.3568 |
| GPT OSS 120B | 2000 | 0.2875 | 0.2438-0.3311 |
| GPT5 nano (high reasoning) | 2000 | 0.1855 | 0.1541-0.2168 |
| GPT5 nano (low reasoning) | 2000 | 0.1796 | 0.1487-0.211 |

**Table 3b:** Agreement analysis of models assessed versus nurse-assigned classifications based on ESI within one ESI level, including exact matches.

| **Model** | **N** | **Agreement within +/- 1 ESI (%)** | **95% CI** |
| --- | --- | --- | --- |
| GPT5.1 (high reasoning) | 16,107 | 90.46 | 90.01-90.91 |
| Stepwise approach (GPT5.1 high reasoning) | 2,000 | 97.85 | 97.20-98.45 |
| Council of Models | 2,000 | 90.4 | 89.1-91.65 |
| GPT5.1 (no reasoning) | 2,000 | 92.75 | 91.6-93.85 |
| Claude Opus 4.5 | 2,000 | 92.7 | 91.55-93.8 |
| GPT5.1 (high reasoning) | 2,000 | 89.45 | 88.1-90.8 |
| Gemini 3 Pro preview | 2,000 | 89.25 | 87.9-90.6 |
| GPT5 Pro (high reasoning) | 2,000 | 87.1 | 85.65-88.55 |
| Mistral Large 3 | 2,000 | 86.8 | 85.3-88.25 |
| GPT5.2 (xhigh reasoning) | 2,000 | 83.8 | 82.2-85.4 |
| Kimi K2 (thinking) | 2,000 | 82.65 | 81.00-84.30 |
| DeepSeek V3.2 | 2,000 | 83.65 | 82.0-85.25 |
| Llama 4 Maverick | 2,000 | 90.7 | 89.45-91.95 |
| Grok 4 | 2,000 | 76.05 | 74.2-77.95 |
| GPT OSS 120B | 2,000 | 82.45 | 80.75-84.05 |
| GPT5 nano (high reasoning) | 2,000 | 62.45 | 60.35-64.60 |
| GPT5 nano (low reasoning) | 2,000 | 62.25 | 60.2-64.4 |

Performance was not uniformly distributed across ESI levels. Confusion matrix analyses and error-distance distributions (Figure 2a, 2b, 2c) revealed several patterns. First, ESI 3 and ESI 4, which together constituted 75.4% of the cohort, were frequently confused across configurations. The Stepwise approach achieved 67.3 % exact agreement for ESI 3 and 89.0 % for ESI 4, whereas the median configuration achieved 42.5 % and 42.7 %, respectively. Second, ESI 2 was over-predicted by the majority of models, consistent with a systematic over-triage pattern. Lastly, only one ESI 1 case was present in the 2,000 cases assessed, which precluded a per-level analysis for the most urgent triage category.

**Figure 2a:**
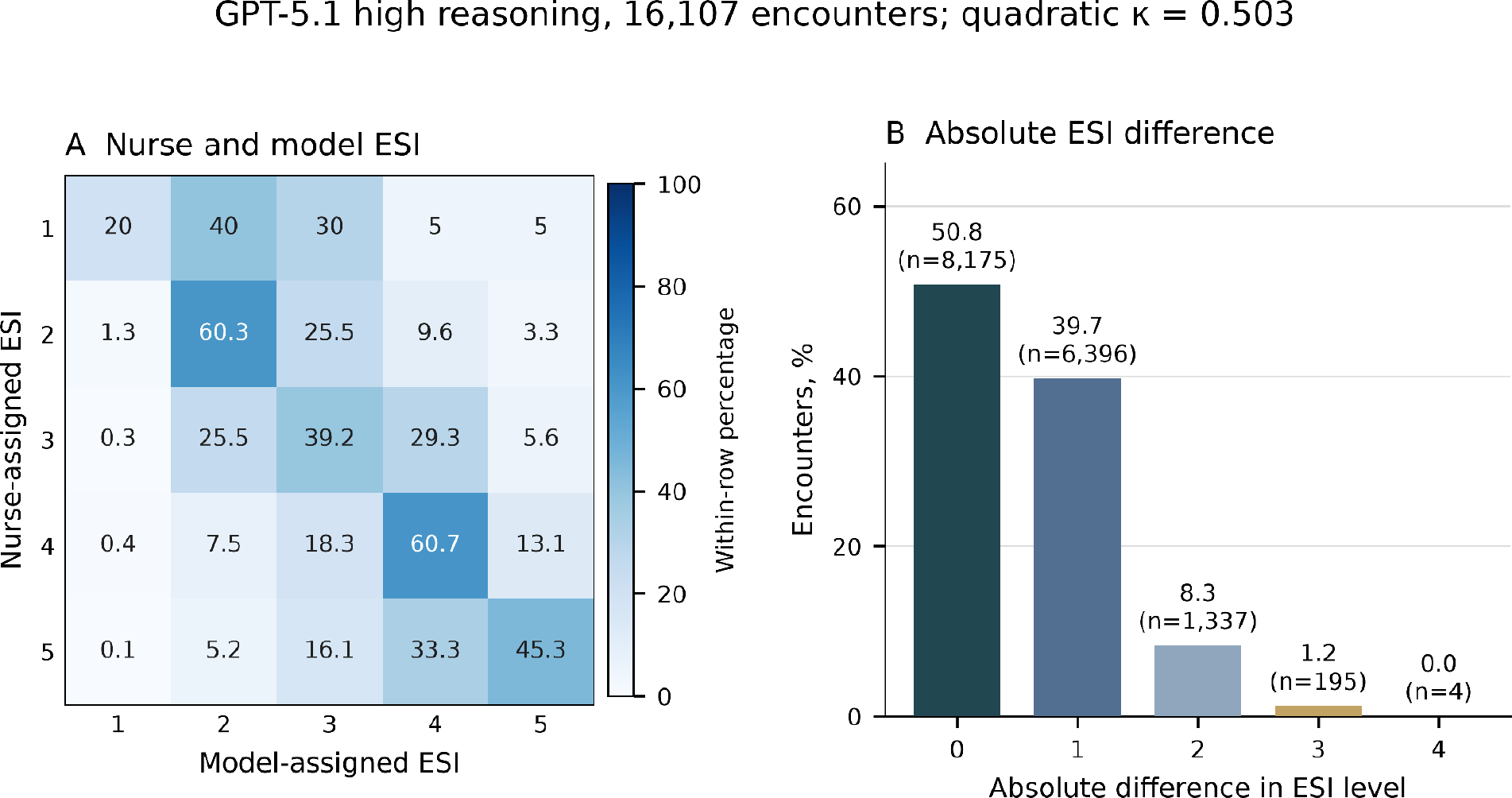
Agreement analysis and error-distance distribution of the full cohort analysis based on GPT 5.1 with high reasoning. ESI: Emergency Severity Index.

**Figure 2b:**
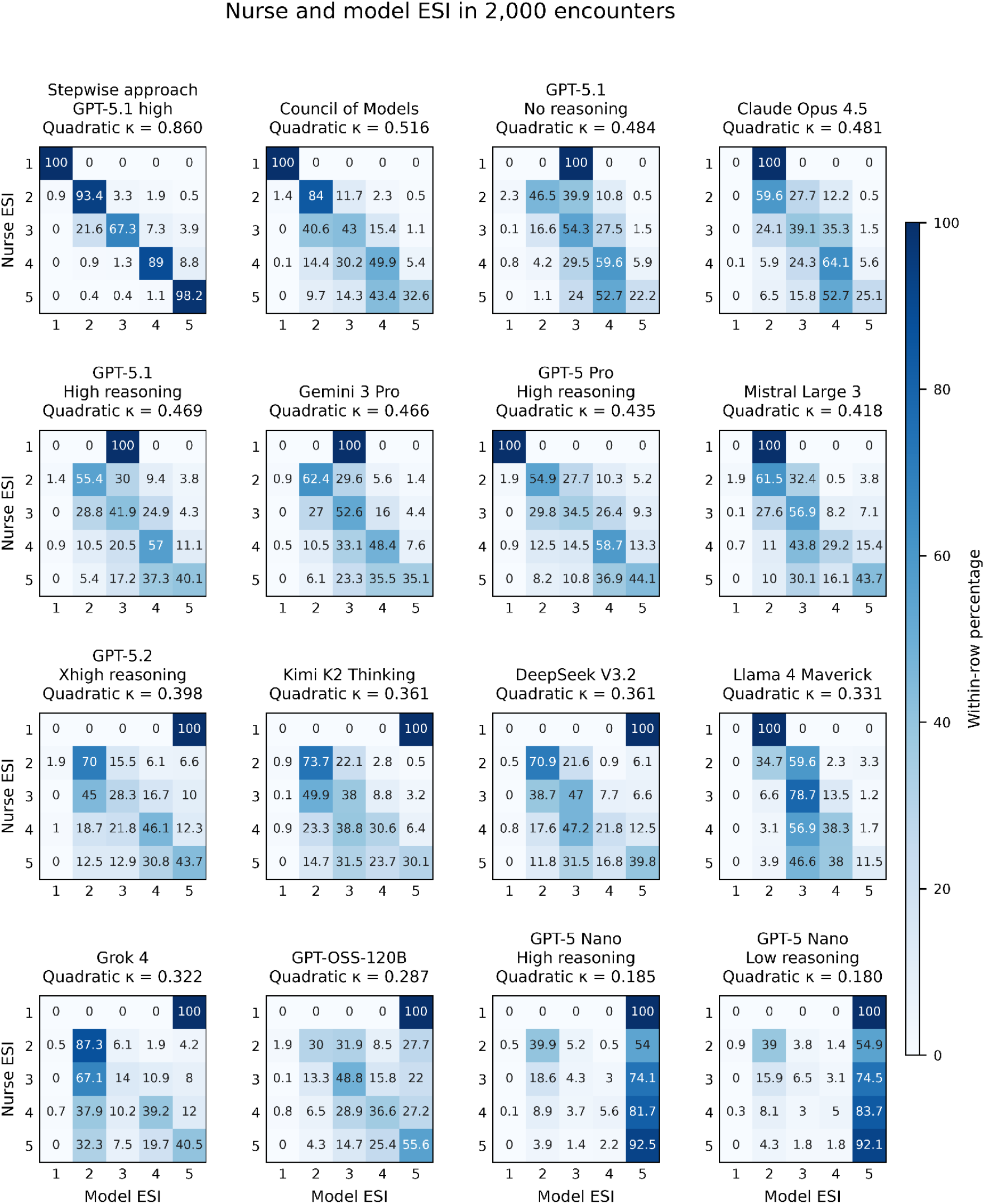
Agreement analysis of the sub cohort analyses based on 16 model configurations. ESI: Emergency Severity Index.

**Figure 2c:**
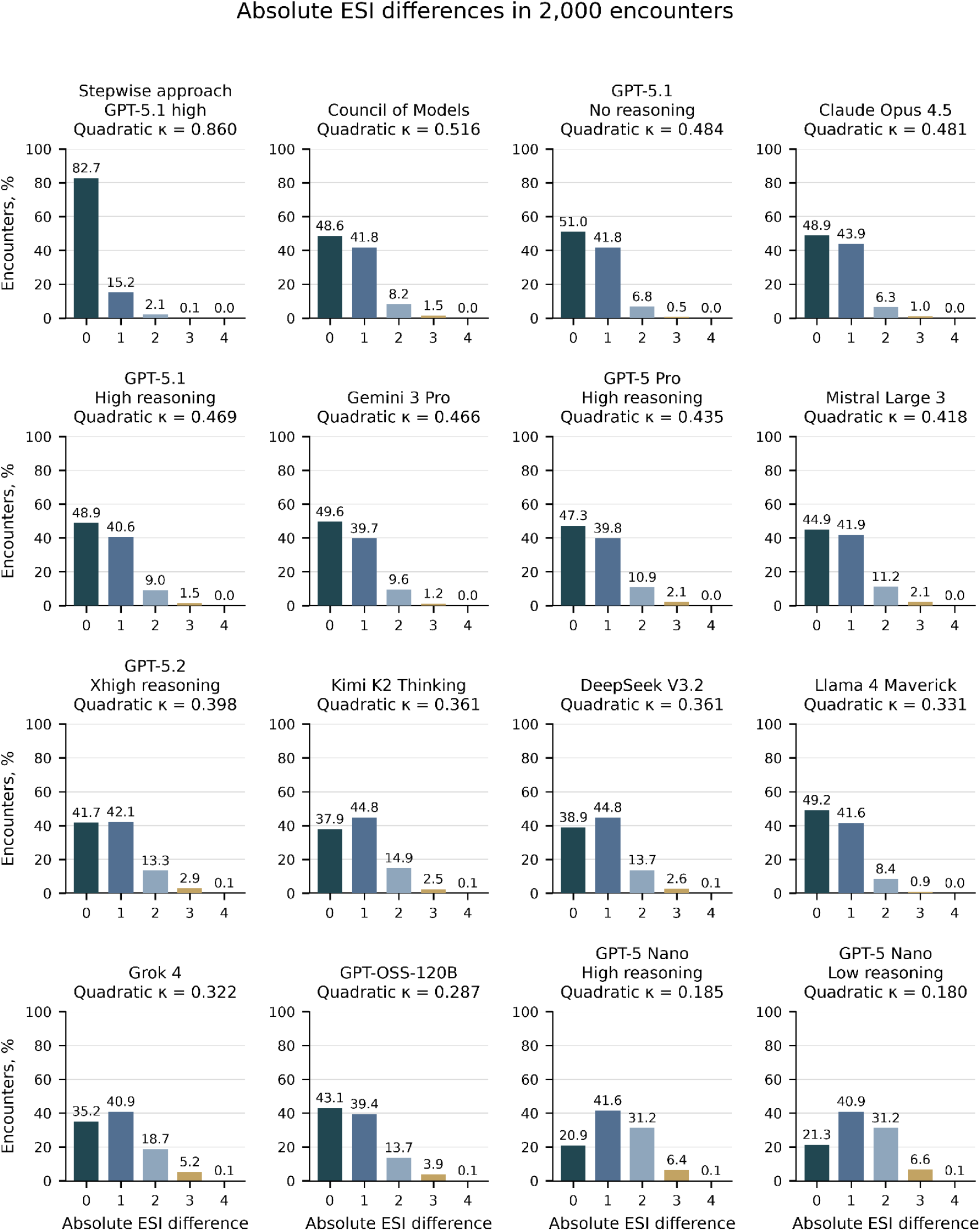
Error-distance distribution of the sub cohort analyses based on 16 model configurations. ESI: Emergency Severity Index.

### Extent of deviation on ESI level

Although a high level of agreement between nursing staff assigned ESI levels and the models assessed is desirable, the extent and direction of any deviation is relevant, too. Both types of misclassification differed substantially across models. Over-triage ranged from 9.20 % for the Stepwise approach to 52.50 % for K2 (Table 4). The Stepwise approach had 8.10 % under-triage and a mean signed deviation of 0.0015 ESI levels. The Council of Models had 41.70 % over-triage and 9.70 % under-triage, while both GPT 5 Nano variants assigned lower acuity than nurses in 66.25 % and 67.25 % of encounters, respectively. Twelve of 16 configurations had a negative mean signed deviation (Table 4). The Stepwise approach assigned ESI 3–5 to 12/214 nurse ESI 1–2 encounters (5.61%; 12/2,000 [0.60%] overall); five of these disagreements involved at least two levels. Across all nurse ESI levels, 34 Stepwise assignments were lower acuity by at least two levels. In the full cohort, standard GPT 5.1 assigned lower acuity to 16/20 nurse ESI 1 encounters and ESI 3–5 to 744/1,937 nurse ESI 1–2 encounters (38.41%).

**Table 4:** Extent of directional disagreement and mean deviation from nurse-assigned acuity classifications based on ESI. Negative signed differences indicate higher model-assigned acuity; positive differences indicate lower acuity.

| Model | N | Over-triage<br>($GPT_{ESI} < Nurse_{ESI}$) (%) | 95% CI | Under-triage<br>($GPT_{ESI} > Nurse_{ESI}$) (%) | 95% CI | Mean deviation<br>($GPT_{ESI} - Nurse_{ESI}$) | 95% CI |
| --- | --- | --- | --- | --- | --- | --- | --- |
| GPT5.1<br>(high reasoning) | 16,107 | 26.67 | 25.98-27.34 | 22.58 | 21.92-23.22 | -0.0675 | -0.0816-<br>-0.053 |
| Stepwise approach<br>(GPT5.1 high reasoning) | 2,000 | 9.20 | 8.00-10.50 | 8.10 | 6.90-9.30 | 0.0015 | -0.0200-0.0225 |
| Council of Models | 2,000 | 41.7 | 39.55-43.85 | 9.7 | 8.4-11 | -0.4155 | -0.4525-<br>-0.3795 |
| GPT5.1<br>(no reasoning) | 2,000 | 30.5 | 28.55-32.5 | 18.5 | 16.8-20.2 | -0.1605 | -0.1975-<br>-0.125 |
| Claude Opus 4.5 | 2,000 | 31 | 29-33.05 | 20.15 | 18.4-21.95 | -0.1525 | -0.1915-<br>-0.1155 |
| GPT5.1<br>(high reasoning) | 2,000 | 31.4 | 29.4-33.45 | 19.75 | 18-21.5 | -0.168 | -0.2095-<br>-0.1275 |
| Gemini 3 Pro preview | 2,000 | 36 | 33.95-38.15 | 14.45 | 12.95-16 | -0.283 | -0.3225-<br>-0.2445 |

| Model | N | Over-triage<br>(GPT<sub>ESI</sub> <<br>Nurse<sub>ESI</sub>) (%) | 95% CI | Under-triage<br>(GPT<sub>ESI</sub> ><br>Nurse<sub>ESI</sub>) (%) | 95% CI | Mean<br>deviation<br>(GPT<sub>ESI</sub> -<br>Nurse<sub>ESI</sub>) | 95% CI |
| --- | --- | --- | --- | --- | --- | --- | --- |
| GPT5 Pro<br>(high<br>reasoning) | 2,000 | 29.75 | 27.8-31.75 | 22.95 | 21.1-24.75 | -0.1045 | -0.1485-<br>-0.0615 |
| Mistral<br>Large 3 | 2,000 | 39.55 | 37.45-41.6<br>5 | 15.55 | 14-17.15 | -0.3225 | -0.3645-<br>-0.2805 |
| GPT5.2<br>(xhigh<br>reasoning) | 2,000 | 40.65 | 38.5-42.8 | 17.65 | 16-19.3 | -0.3035 | -0.35-<br>-0.2585 |
| Kimi K2<br>(thinking) | 2,000 | 52.5 | 50.35-54.7 | 9.65 | 8.40-10.95 | -0.5925 | -0.6335-<br>-0.5505 |
| DeepSeek<br>V3.2 | 2,000 | 47.9 | 45.75-50.1 | 13.2 | 11.75-14.7 | -0.4575 | -0.502-<br>-0.414 |
| Llama 4<br>Maverick | 2,000 | 37.75 | 35.65-39.9<br>5 | 13.1 | 11.6-14.6 | -0.3205 | -0.358-<br>-0.284 |
| Grok 4 | 2,000 | 51.9 | 49.8-54.05 | 12.95 | 11.5-14.4 | -0.598 | -0.6475-<br>-0.549 |
| GPT OSS<br>120B | 2,000 | 25.25 | 23.4-27.2 | 31.7 | 29.7-33.7 | 0.1515 | 0.1025-0.2 |
| GPT5<br>nano (high<br>reasoning) | 2,000 | 12.85 | 11.45-14.3<br>5 | 66.25 | 64.2-68.3 | 0.878 | 0.824-0.92<br>95 |
| GPT5<br>nano (low<br>reasoning) | 2,000 | 11.45 | 10.05-12.8<br>5 | 67.25 | 65.15-69.3 | 0.907 | 0.8545-0.9<br>6 |

### Sectoral assignment agreement

Agreement on the care sector assignment (i.e. ED or UCP) was low across all models (Table 5). Unweighted κ values ranged from 0.088 (GPT 5 Nano) to 0.304 (Council of Models), corresponding to slight-to-fair agreement. Misallocations to UCP (i.e. model decision to allocate patients to the UCP when the nursing-assigned allocation was the ED) ranged from 5.71% (Kimi K2) to 75.38% (GPT 5 Nano, reasoning low). This type of misclassification occurred most frequently in the GPT 5 Nano models with similar frequencies under the two evaluated reasoning settings (Table 5). Vice versa, Misallocations to ED (i.e. model decision to allocate patients to the ED when the nursing-assigned allocation was to the UCP) ranged from 10.87% (GPT 5 Nano, reasoning low) to 83.94% (Kimi K2) (Table 5).

**Table 5:**
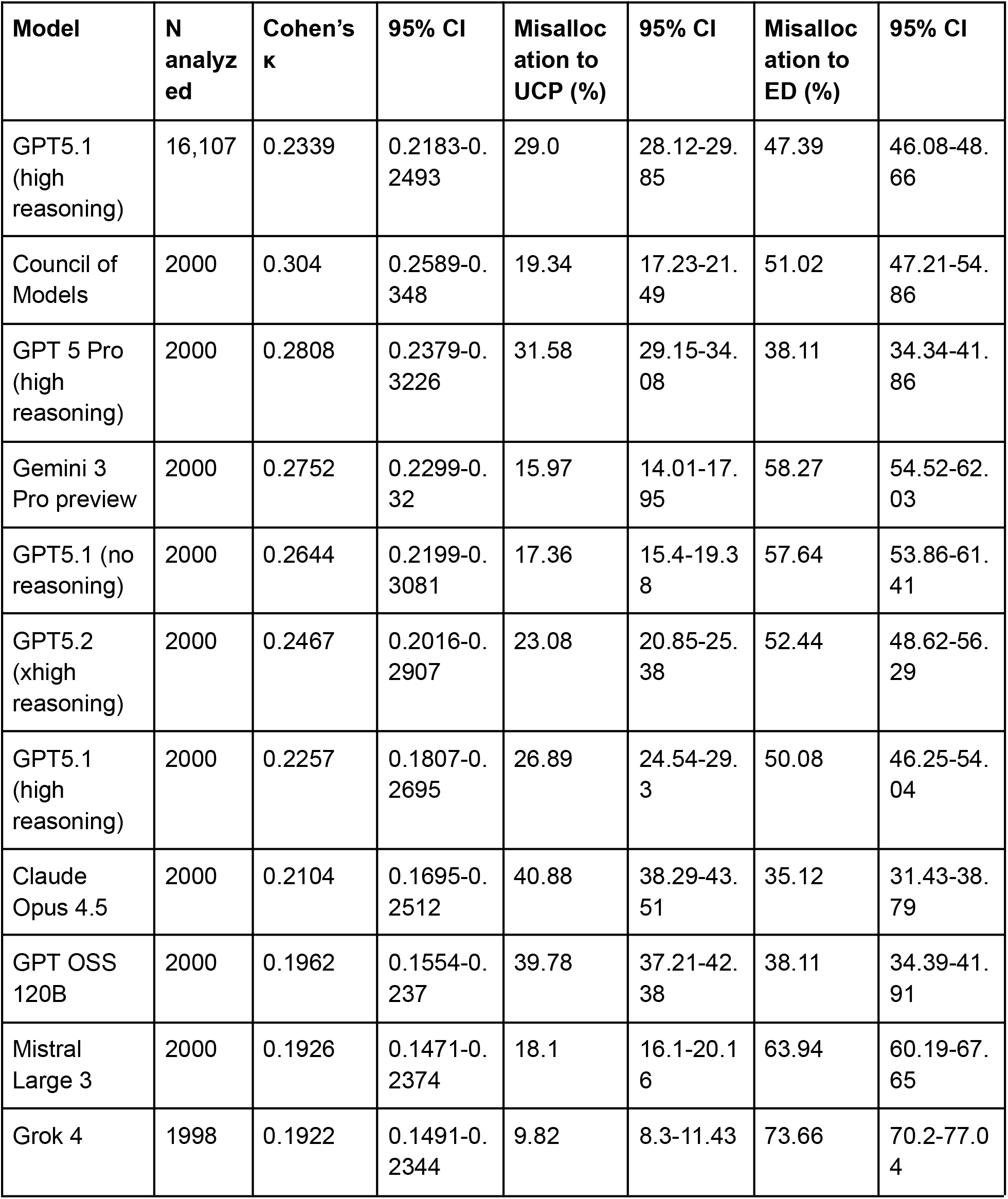
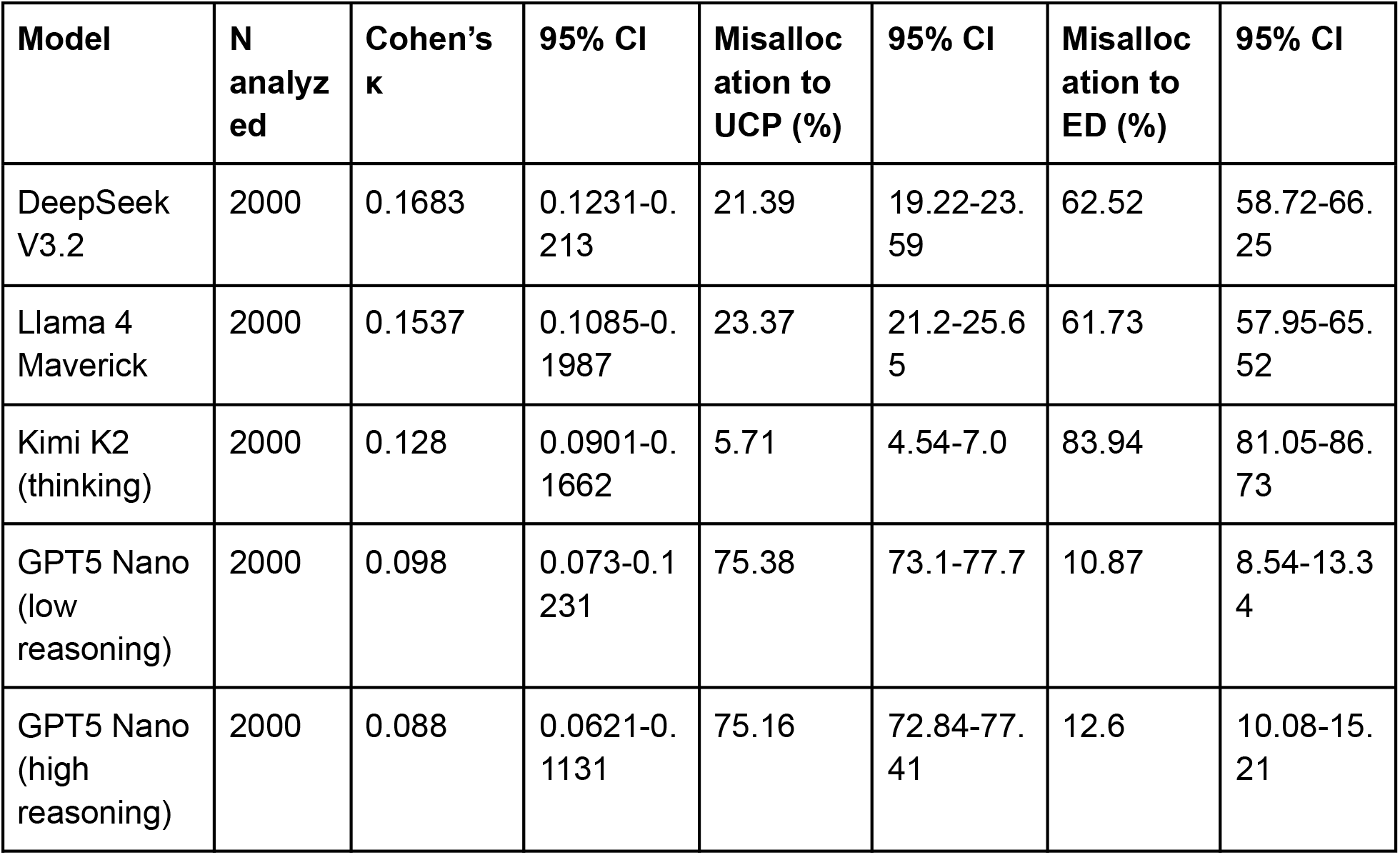
Agreement analysis of sectoral allocation between nursing-assigned referral destination and model decision. Misallocation to UCP: Model decision to allocate patients to the UCP when the nursing-assigned allocation was ED. Misallocation to ED: Model decision to allocate patients to the ED when the nursing-assigned allocation was UCP.

| **Model** | **N analyzed** | **Cohen's $\kappa$** | **95% CI** | **Misallocation to UCP (%)** | **95% CI** | **Misallocation to ED (%)** | **95% CI** |
| --- | --- | --- | --- | --- | --- | --- | --- |
| GPT5.1 (high reasoning) | 16,107 | 0.2339 | 0.2183-0.2493 | 29.0 | 28.12-29.85 | 47.39 | 46.08-48.66 |
| Council of Models | 2000 | 0.304 | 0.2589-0.348 | 19.34 | 17.23-21.49 | 51.02 | 47.21-54.86 |
| GPT 5 Pro (high reasoning) | 2000 | 0.2808 | 0.2379-0.3226 | 31.58 | 29.15-34.08 | 38.11 | 34.34-41.86 |
| Gemini 3 Pro preview | 2000 | 0.2752 | 0.2299-0.32 | 15.97 | 14.01-17.95 | 58.27 | 54.52-62.03 |
| GPT5.1 (no reasoning) | 2000 | 0.2644 | 0.2199-0.3081 | 17.36 | 15.4-19.38 | 57.64 | 53.86-61.41 |
| GPT5.2 (xhigh reasoning) | 2000 | 0.2467 | 0.2016-0.2907 | 23.08 | 20.85-25.38 | 52.44 | 48.62-56.29 |
| GPT5.1 (high reasoning) | 2000 | 0.2257 | 0.1807-0.2695 | 26.89 | 24.54-29.3 | 50.08 | 46.25-54.04 |
| Claude Opus 4.5 | 2000 | 0.2104 | 0.1695-0.2512 | 40.88 | 38.29-43.51 | 35.12 | 31.43-38.79 |
| GPT OSS 120B | 2000 | 0.1962 | 0.1554-0.237 | 39.78 | 37.21-42.38 | 38.11 | 34.39-41.91 |
| Mistral Large 3 | 2000 | 0.1926 | 0.1471-0.2374 | 18.1 | 16.1-20.16 | 63.94 | 60.19-67.65 |
| Grok 4 | 1998 | 0.1922 | 0.1491-0.2344 | 9.82 | 8.3-11.43 | 73.66 | 70.2-77.04 |
| DeepSeek V3.2 | 2000 | 0.1683 | 0.1231-0.213 | 21.39 | 19.22-23.59 | 62.52 | 58.72-66.25 |
| Llama 4 Maverick | 2000 | 0.1537 | 0.1085-0.1987 | 23.37 | 21.2-25.65 | 61.73 | 57.95-65.52 |
| Kimi K2 (thinking) | 2000 | 0.128 | 0.0901-0.1662 | 5.71 | 4.54-7.0 | 83.94 | 81.05-86.73 |
| GPT5 Nano (low reasoning) | 2000 | 0.098 | 0.073-0.1231 | 75.38 | 73.1-77.7 | 10.87 | 8.54-13.34 |
| GPT5 Nano (high reasoning) | 2000 | 0.088 | 0.0621-0.1131 | 75.16 | 72.84-77.41 | 12.6 | 10.08-15.21 |

### Confidence assessment

All 14 individually evaluated configurations had mean verbalized confidence above exact agreement (Table 6). Mean confidence ranged from 52.1% to 67.9%, while exact ESI agreement ranged from 20.9% to 51.0%, yielding confidence–agreement gaps of +5.2 to +33.7 percentage points. Verbalized confidence distributions for the comparison sample are shown in Annex G. The weighted absolute bin-level gap ranged from 0.099 for Claude Opus 4.5 to 0.357 for GPT 5 Nano with high reasoning effort. Confidence accuracy (CA) ranged from −0.0489 to 0.0234, corresponding to mean confidence differences between exact matches and mismatches of −4.89 to 2.34 percentage points. Confidence–agreement plots, adapted from reliability diagrams ^48^, are shown for the comparison sample in Annex H. Corresponding analyses for the full cohort of 16,107 encounters evaluated using GPT 5.1 with high reasoning effort are presented in Annex I. The Stepwise approach was excluded from confidence analyses because its workflow did not provide a directly comparable final encounter-level confidence score.

**Table 6:**
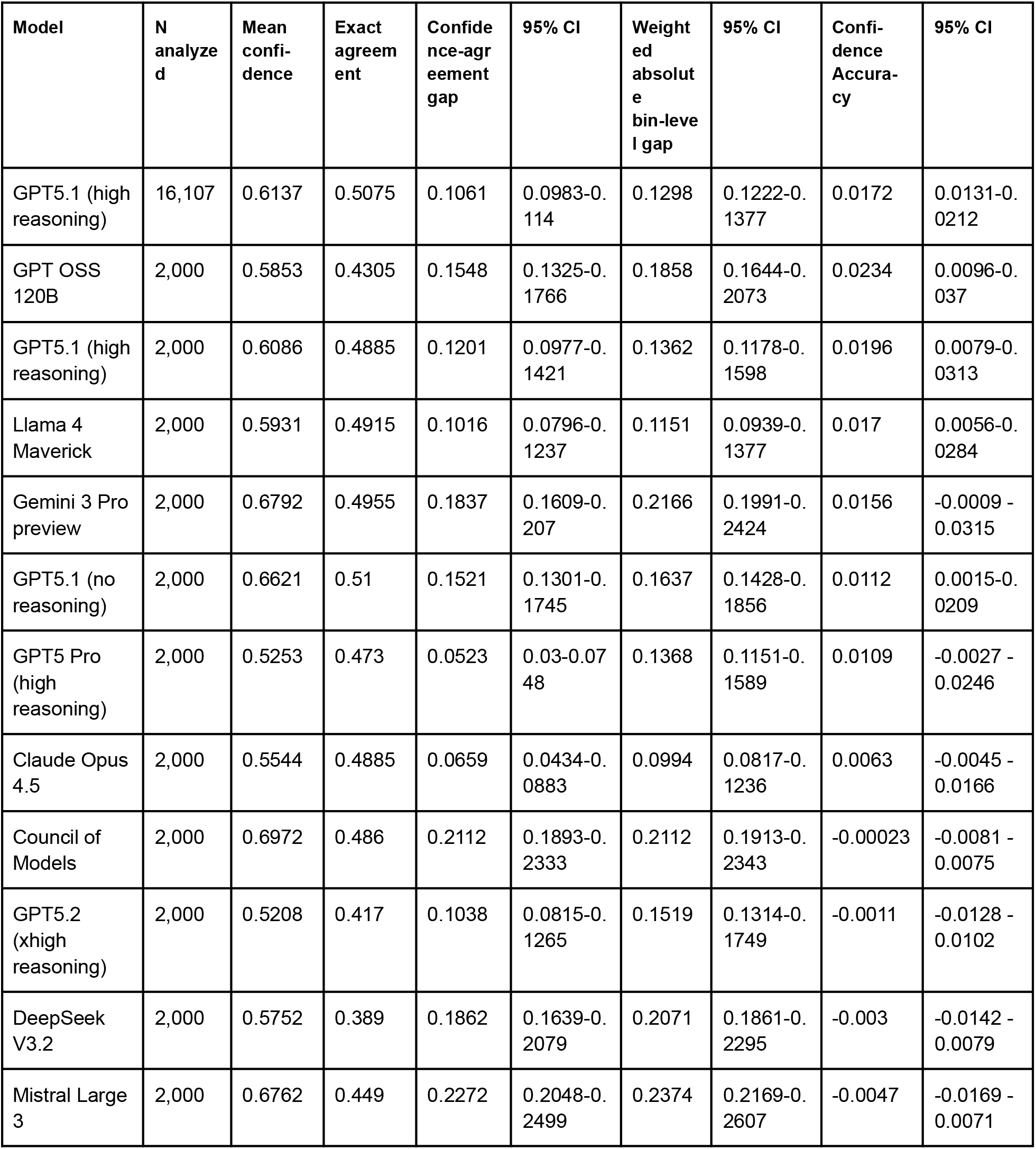
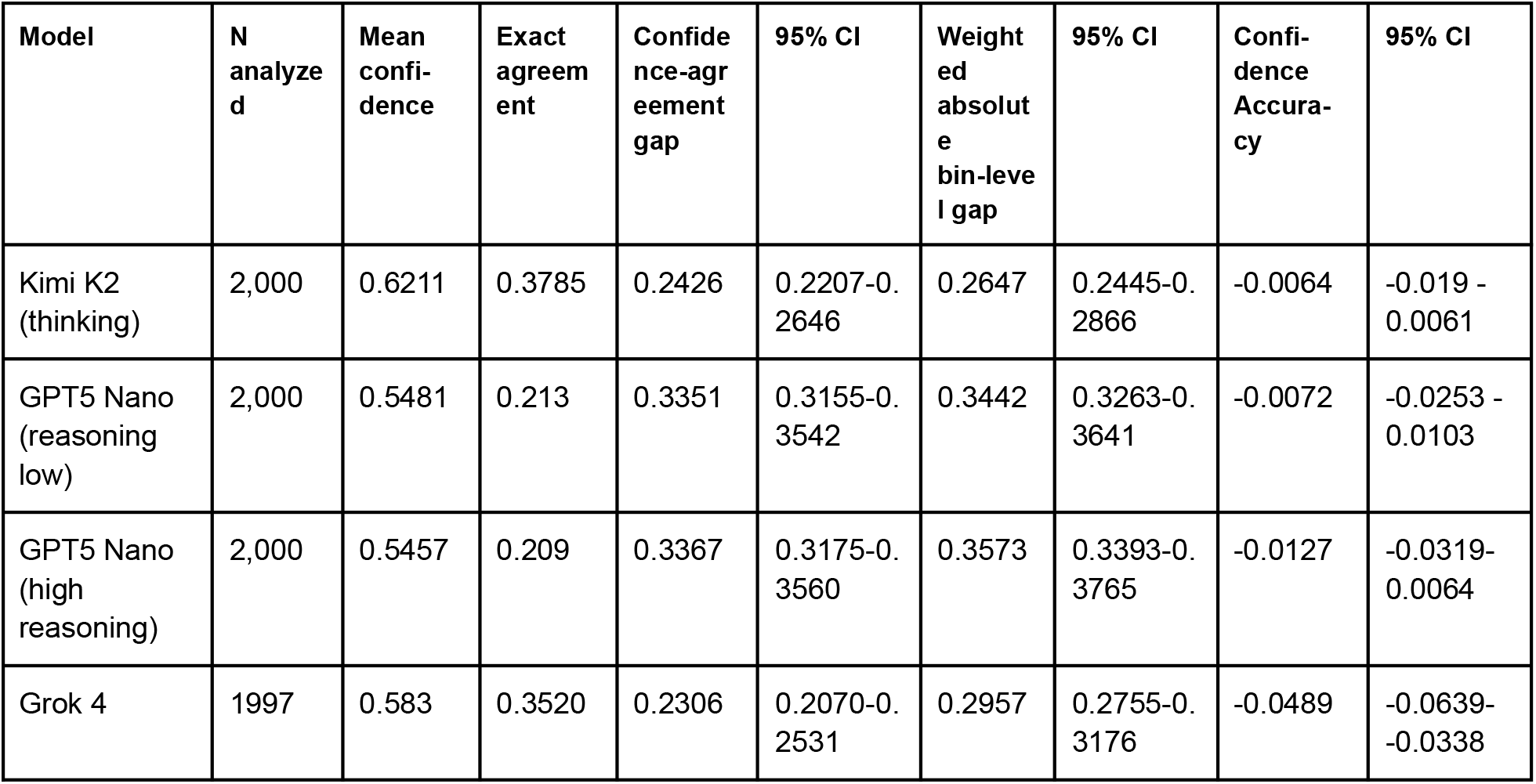
Descriptive comparison of model-reported verbalized confidence with exact agreement and nurse-assigned ESI. Confidence scores were expressed on a 0–1 scale. The confidence–agreement gap is mean confidence minus exact agreement. The weighted absolute bin-level gap is the sum of absolute differences between mean confidence and exact agreement within each confidence bin, weighted by the proportion of encounters in that bin. Confidence accuracy (CA) is mean confidence among exact ESI matches minus mean confidence among ESI mismatches. These measures describe confidence in relation to nurse agreement.

### Consistency of model output

In the reproducibility experiment, GPT 5.1 (high reasoning effort) was queried 100 times with identical input for each of 100 encounters, yielding 10,000 classifications. Multi-rater reliability across runs was substantial (Fleiss’ κ = 0.716), and the median encounter received the same modal ESI classification in 95 of 100 runs. However, consistency varied substantially across encounters. While 61 % of encounters achieved high consistency (modal frequency ≥90 %, including 30 encounters with identical ESI outputs across all observed runs), 23 % of encounters received their most common ESI classification in fewer than 70 of 100 runs (Figure 3).

**Figure 3:**
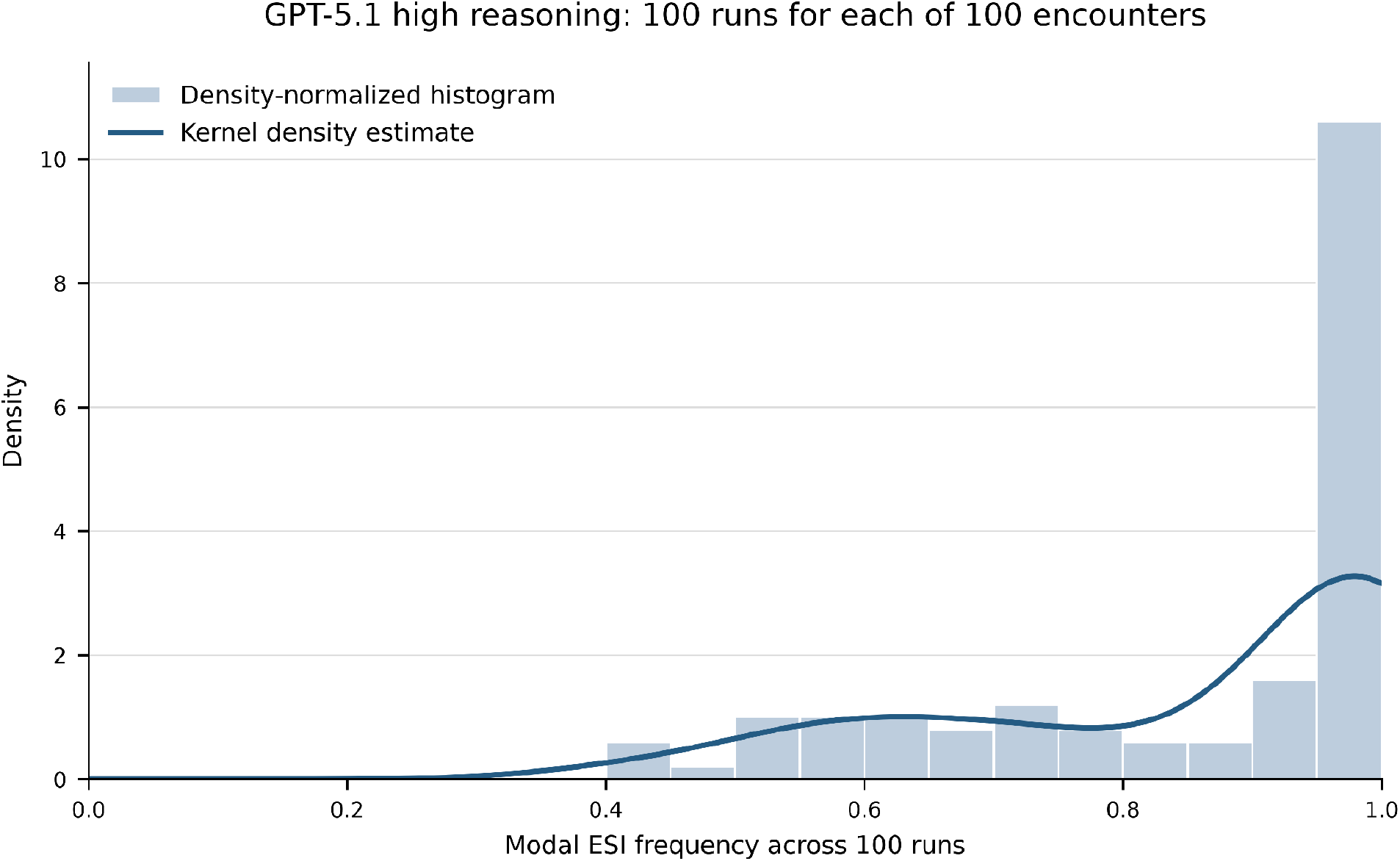
Consistency of acuity assessment based on 100 repeated runs of a subsample of 100 encounters. Consistency is the frequency of the modal ESI assignment.

Notably, modal aggregation over multiple runs did not improve mean exact agreement. Exact-match agreement with the triage nursés ESI level was 41.11 % averaged across individual runs and 41.00 % for the modal assignment over all 100 runs, a paired difference of −0.11 percentage points (95% CI −3.79 to 3.66).

## Discussion

### Primary Findings

In this systematic evaluation of one full cohort assessment of 16,107 encounters by GPT 5.1 with high reasoning effort and 16 sub-cohort analyses by a broad set of contemporary LLM setups for emergency department triage, we found that most evaluated configurations i) achieve only slight-to-moderate agreement with nurse-assigned ESI levels, ii) express mean confidence above exact agreement in all 14 individual configurations, and iii) in the GPT 5.1 reproducibility experiment, produce inconsistent outputs for a substantial minority of encounters (i.e. 23% received their most frequent ESI classification in < 70/100 runs). The Stepwise approach achieved the highest agreement (κ_qw = 0.860; exact agreement 82.70%), comparable to nurse-to-nurse reliability from separate studies ^10,49^, although design and analytical approach warrant a careful interpretation and cannot establish human equivalence.

#### Model performance in the context of nurse agreement

A multicenter study reported weighted κ of 0.70–0.80 for written scenarios and 0.69–0.87 for actual triages ^10,49^. The Stepwise approach achieved κ_qw = 0.860 and the next-best approach (Council of Models) achieved κ_qw = 0.516, which remains well below these measures. Yet, these results warrant a careful interpretation, because case mix, available information and reference standards differ. Likewise, several prior studies have reported higher LLM agreement for triage tasks, while direct comparison remains limited by differences in datasets, triage systems, outcome definitions, and the number of models evaluated ^27,28,50–52^. Our study, however, evaluates more than a dozen contemporary model configurations on a common real-world comparison sample and the wide range of observed κ_qw values (0.180 to 0.860) underscores the risk of generalising from single-model studies: the choice of model and prompting strategy greatly influences performance, and results from one model cannot be extrapolated to others.

### Secondary Findings

#### Misclassification and potential implications

Our findings warrant a careful discussion of the applicability of LLMs for clinical decision making. Most discrepancies in classifications occurred between adjacent ESI levels and especially between ESI 3 and ESI 4, which highlights a level of ambiguity between moderate acuity levels that is well known in clinical practice ^53^ ^54^. Yet, the low consistency of assessments for a relevant proportion of cases raises the question of whether this ambiguity is more rooted in the probabilistic nature of the models assessed, or whether it truly reflects the clinical challenge of assessing acuity ^31^. In our particular setting, the sectoral assignment is a crucial step for downstream resource allocation and all models assessed performed poorly. Concretely, over-triage could affect resource use if these configurations were deployed. Conversely, both GPT Nano variants assigned lower acuity than nurses in approximately two thirds of encounters. The Stepwise approach had 8.10 % lower-acuity assignments, including 12 of 214 nurse ESI 1–2 encounters assigned ESI 3–5, which warrants clinical review.

#### Relevance of prompting strategy and model characteristics

Prior work has evaluated prompt strategies for medical question answering ^55^, chain-of-thought reasoning across benchmark tasks ^56^, and an LLM pipeline for cardiovascular risk-score calculation ^57^. Our comparison included direct ESI classification and a Stepwise workflow combining structured intermediate model assessments with deterministic ESI mapping. This approach yielded an unexpected, yet relevant additional finding. The structured ESI Stepwise approach outperformed all other configurations by a relevant margin, including configurations using the same underlying model (GPT 5.1) with different prompting strategies. On the same 2,000 encounters, κ_qw was 0.469 for standard GPT 5.1 high reasoning and 0.860 for the Stepwise approach, a paired difference of 0.391 (95% CI 0.354–0.429). Exact agreement differed by 33.85 percentage points (95% CI 31.40–36.35). This comparison evaluates complete workflows and does not isolate the effects of prompting or deterministic mapping.

#### Verbalized confidence measures for clinical decision support

Mean verbalized confidence exceeded exact ESI agreement in all 14 individual configurations, with confidence–agreement gaps ranging from 5.2 to 33.7 percentage points. Confidence accuracy (CA) indicated small differences in mean confidence between matching and nonmatching ESI assignments, ranging from −4.89 to 2.34 percentage points. In clinical practice, confidence scores are sometimes displayed alongside decision-support recommendations to help clinicians calibrate their trust ^58^. However, the usefulness of the elicited certainty score as a reliability signal for clinical triage remains unestablished.

#### Reproducibility and implications for clinical safety

The observation that 23 % of encounters had modal ESI frequency below 70% across 100 repeated calls demonstrates output variability in this clinical triage dataset. For these encounters, the repeated classifications span multiple acuity levels that could lead to different care pathways if used clinically. This variability requires prospective evaluation at the critical decision point of medical triage.

The balance between different ESI levels varied across runs. This could reflect clinical ambiguity, prompt interpretation or model stochasticity. Yet, without matched expert review and complete request settings, these explanations cannot be distinguished.

#### Sectoral assignment Agreement

Agreement on the care sector assignment (i.e. UCP or ED) was low across the evaluated configurations, with no configuration exceeding κ = 0.304. A potential explanation is that referral assignments in the clinical context at hand are not yet standardized, scarcely represented in the training data of the models and are contingent upon local institutional protocols, resource availability, and institution-specific clinical knowledge (e.g., particular presenting complaints are routed to the ED because of local resource constraints), which may not be reflected in the patient data provided to the model. This finding suggests that referral assignment may require institution-specific fine-tuning or retrieval-augmented approaches that incorporate local routing protocols.

### The usability of LLM-based clinical decision support systems in a real-world care setting

The deployment of LLM-based and other artificial intelligence solutions in a real-world healthcare context often faces tension between the technological promise and the applicability in a real-world setting. This work aimed at exploring this applicability using routine recorded data alongside prior in silico evaluations ^27–31^ and identified distinct challenges that warrant a discussion.

Firstly, the availability of high quality triage data is an important feature of a useful CDSS. Although not systematically assessed for this work, free-text entries show a high proportion of typographical errors and the use of not established abbreviations or jargon. Likewise, a relevant proportion of vital sign categories are not filled and are hence not available for a CDSS. Yet, these challenges, ranging from incomplete to typographically inaccurate triage data, are not unique to our study, but typical for a broad spectrum of emergency care settings ^59–61^ and robust CDSS solutions need to account for these shortcomings.

Secondly, regulatory requirements for both the use of triage data and resulting outputs from a CDSS apply. While anonymous data do not fall under the European General Data Protection Regulation (GDPR), linkage to downstream care data would require an appropriate legal basis and institutional authorization. These arrangements were not established for this analysis. Similarly, LLM-based systems intended for a medical purpose may be subject to the European Medical Devices Regulation (MDR). This applies, for example, to systems that assist in diagnoses, calculate risks, or generate therapeutic recommendations, all applying to the use of a CDSS for triage and referral decisions. AI-based medical devices are furthermore subject to the EU AI Regulation 2024/1689/EU. This regulation imposes particular obligations on providers and operators of AI systems. Applicable high-risk AI requirements can include mandatory human oversight and data-quality obligations. In general, regulatory compliance and external-provider processing arrangements would require an assessment before clinical deployment.

Thirdly, a genuine assessment of triage performance needs to reflect relevant outcomes that extend beyond agreement in the triage and referral assessment. The very nature of triage is error-prone, as categorical decisions need to be made with a significant level of uncertainty. Studies using different reference standards have reported substantial mistriage, with variation across demographic and socioeconomic strata ^62,63^. As comparing a CDSS merely to a nursing staff standard at the time of triage ignores this complexity, we deem a comprehensive performance assessment necessary that includes relevant outcome data reflecting the entire episode of care (e.g. triage-to-discharge) that would allow an estimation of the clinical relevance of both mistriage and sectoral misfit. Yet, these data were not available for our analysis because downstream records could not be linked under the available study arrangements, as outlined above.

### Limitations

This work is based on an uncurated real-world data set and is prone to several biases that warrant a careful interpretation of our results.

Firstly, only walk-in patients are assessed with the CDSS at the time of arrival and hence, the data set has a relevant selection bias against critically ill patients who are often admitted through EMS.

Secondly, documentation fatigue is likely to contribute to a detection bias, as nursing staff might not document all data that are relevant for their assessment of severity and assignment of the care sector. The nurse assessment is therefore an unmatched reference rather than an independently adjudicated gold standard.

Thirdly, due to the operational complexity of the UCP on premise of a large academic medical center, the UCP is unavailable at late night hours (11 pm through 8 am), which limits the nursing staff’s ability to freely choose the appropriate care sector at night. Recorded overnight timestamps occurred in 1,041/2,000 comparison encounters (52.05%), versus 1,832/16,107 encounters overall (11.37%).

Fourthly, nursing staff could be influenced in their decision-making by the suggestions from the CDSS (TriageClient). Although the final decision is always made by the nursing staff, a potential influence cannot be ruled out.

In summary, we deem the data set a robust foundation for this analysis, as it highlights the challenges of using uncurated real-world data for clinical decision making at a highly relevant decision point in emergency and acute care settings.

### Conclusion

In this large-scale real-world evaluation, most evaluated configurations showed only modest agreement with nurse-assigned triage decisions. Mean confidence exceeded exact agreement in all 14 individual configurations; output variability was assessed only for GPT 5.1. The Stepwise approach achieved the highest agreement. Reproducibility experiments showed variation across repeated classifications. The unmatched reference and limited representation of critically ill patients preclude conclusions about clinical safety or human equivalence. While LLMs hold potential as supportive tools in emergency care, their current performance characteristics do not yet support autonomous or safety-critical deployment in clinical triage environments, and further and prospective evaluations are warranted.

## Authorship Contributions

LB developed the study design, defined research questions, performed AI Inference and statistical analysis, wrote the draft of the manuscript and should be considered first author. AH contributed to the formal analysis. MG, TR and MS provided methodological guidance and contributed to the formal analysis. FPH contributed to the development of the study design, provided the data used for analysis and contributed to draft of the manuscript. TU contributed to the development of the study design. HJB provided clinical supervision and guidance for the clinical context of the study. AM provided methodological guidance and the technical context of the study and should be considered last author. JM developed the study design, defined research questions, performed AI Inference and statistical analysis, wrote the draft of the manuscript and should be considered last author. All authors contributed to revising the manuscript prior to submission.

## Data Availability Statement

The datasets generated and/or analysed during the current study are not publicly available due to institutional data protection requirements, but are available from the corresponding author on reasonable request.

## Code Availability Statement

The analysis code is available from the corresponding author on reasonable request. Model prompts and the deterministic Stepwise decision rules are provided in Annex D.

