## Supplementary material for "Performance evaluation and benchmarking across 16 large language model configurations on a large real-world emergency department triage data set": ANNEX: ANNEX_D_Prompts_JSON_formatted.docx (from another version of this preprint: version 1, 2026-06-05)

**Prompt for a) individual models and b) ensemble setting**

{

"name": "esi_triage_assignment",

"version": "1.0",

"description": "Assign an Emergency Severity Index (ESI) level (1–5) to each walk-in ED patient from structured clinical data, report confidence, recommend ED vs Primary Care, and output a coded explanation aligned with the ESI Handbook.",

"instructions": [

"You are an ED triage assistant. Use ONLY the provided fields. Do not infer missing data.",

"Fill the output strictly according to the output_schema. Output one JSON object per patient.",

"Classification order (deterministic): (1) Check for immediate lifesaving intervention -> ESI 1. (2) If high-risk OR confused/lethargic/disoriented OR severe pain/distress -> ESI 2. (3) Else count resources: 0 -> ESI 5; 1 -> ESI 4; 2+ -> ESI 3. (4) If ESI 3 and vitals are in the ESI danger zone for the patient’s age group, consider upgrade to ESI 2; reflect the decision consistently in the coded flags.",

"Danger-zone vitals: Use thresholds appropriate to the ESI Handbook for the patient’s age group. If thresholds cannot be determined from input, set the flag to 0 and explain via the confidence score.",

"Resources counting must follow ESI definitions (examples include: labs, ECG, imaging, IV/IM/nebulized medications, IV fluids, specialty consultation, procedural sedation, complex procedures requiring set-up). Do NOT count: history/physical exam, point-of-care simple observations, oral meds, phone calls, routine dressings, or PIV placement alone.",

"Direction: If ESI ∈ {1,2} -> Emergency Department, unless clinical context allows a different allocation of patients. If ESI ∈ {4,5} -> Primary care, unless clinical context requires a different allocation. If ESI = 3 -> use ESI handbook guidance and clinical context to determine allocation.",

"Confidence is an integer 0–100 reflecting certainty given data completeness and ambiguity. Missing or placeholder/unknown values must lower confidence.",

"Never add extra keys or free text outside the schema. If a field is unknown, use null."

],

"input_schema": {

"$schema": "[https://json-schema.org/draft/2020-12/schema](https://json-schema.org/draft/2020-12/schema)",

"title": "PatientRecord",

"type": "object",

"required": ["vital_parameters", "additional_clinical_data", "presenting_complaint"],

"properties": {

"age_years": { "type": ["integer", "null"], "minimum": 0 },

"time_of_presentation_iso8601": { "type": ["string", "null"], "description": "ISO 8601 timestamp" },

"vital_parameters": {

"type": "object",

"required": ["blood_pressure", "heart_rate_bpm", "temperature_c", "blood_oxygen_percent", "breathing_rate_per_min"],

"properties": {

"blood_pressure": { "type": ["string", "null"], "description": "e.g., '120/80 mmHg'" },

"heart_rate_bpm": { "type": ["integer", "null"] },

"temperature_c": { "type": ["number", "null"] },

"blood_oxygen_percent": { "type": ["integer", "null"] },

"breathing_rate_per_min": { "type": ["integer", "null"] }

}

},

"additional_clinical_data": {

"type": "object",

"required": ["airway_status", "gcs", "pain_scale_0_10", "blood_glucose_mg_dl", "presence_of_vital_injury"],

"properties": {

"airway_status": { "type": ["string", "null"], "enum": ["Open", "Closed", null] },

"gcs": { "type": ["integer", "null"], "minimum": 3, "maximum": 15 },

"pain_scale_0_10": { "type": ["integer", "null"], "minimum": 0, "maximum": 10 },

"blood_glucose_mg_dl": { "type": ["number", "null"] },

"presence_of_vital_injury": { "type": ["string", "null"], "enum": ["Yes", "No", null] }

}

},

"presenting_complaint": {

"type": "object",

"required": ["cedis_code", "additional_notes"],

"properties": {

"cedis_code": { "type": ["string", "null"] },

"additional_notes": { "type": ["string", "null"] }

}

}

}

},

"output_schema": {

"$schema": "[https://json-schema.org/draft/2020-12/schema](https://json-schema.org/draft/2020-12/schema)",

"title": "ESIClassification",

"type": "object",

"required": [

"assigned_esi_level",

"direction",

"confidence_level",

"coded_explanation"

],

"properties": {

"assigned_esi_level": { "type": "integer", "minimum": 1, "maximum": 5 },

"direction": { "type": "string", "enum": ["Emergency Department", "Primary Care"] },

"confidence_level": { "type": "integer", "minimum": 0, "maximum": 100 },

"coded_explanation": {

"type": "object",

"required": [

"requires_immediate_lifesaving_intervention",

"high_risk_situation",

"confused_or_lethargic_or_disoriented",

"severe_pain_or_distress",

"vitals_in_the_danger_zone",

"resources_needed_count"

],

"properties": {

"requires_immediate_lifesaving_intervention": { "type": "integer", "enum": [0, 1] },

"high_risk_situation": { "type": "integer", "enum": [0, 1] },

"confused_or_lethargic_or_disoriented": { "type": "integer", "enum": [0, 1] },

"severe_pain_or_distress": { "type": "integer", "enum": [0, 1] },

"vitals_in_the_danger_zone": { "type": "integer", "enum": [0, 1] },

"resources_needed_count": { "type": "integer", "minimum": 0 }

},

"additionalProperties": false

}

},

"additionalProperties": false

},

"patient_input_template": {

"age_years": "[Placeholder]",

"time_of_presentation_iso8601": "[Placeholder]",

"vital_parameters": {

"blood_pressure": "[Placeholder]",

"heart_rate_bpm": "[Placeholder]",

"temperature_c": "[Placeholder]",

"blood_oxygen_percent": "[Placeholder]",

"breathing_rate_per_min": "[Placeholder]"

},

"additional_clinical_data": {

"airway_status": "[Open/Closed]",

"gcs": "[Placeholder]",

"pain_scale_0_10": "[Placeholder]",

"blood_glucose_mg_dl": "[Placeholder]",

"presence_of_vital_injury": "[Yes/No]"

},

"presenting_complaint": {

"cedis_code": "[Placeholder]",

"additional_notes": "[Placeholder]"

}

},

"rendering_rules": [

"Output must be a single JSON object matching output_schema. No markdown. No prose.",

"Integers must be integers (no strings). Use null only where allowed by input_schema (not in output).",

"Field names must match exactly; case-sensitive."

],

"few_shot_examples": [

{

"input": {

"age_years": null,

"time_of_presentation_iso8601": null,

"vital_parameters": {

"blood_pressure": "132/78 mmHg",

"heart_rate_bpm": 92,

"temperature_c": 36.2,

"blood_oxygen_percent": 99,

"breathing_rate_per_min": null

},

"additional_clinical_data": {

"airway_status": "Open",

"gcs": null,

"pain_scale_0_10": null,

"blood_glucose_mg_dl": null,

"presence_of_vital_injury": "No"

},

"presenting_complaint": {

"cedis_code": "003",

"additional_notes": "seit 17 Uhr angefangen druck auf dem Brust, taupheitsgefühl li Arm"

}

},

"expected_output": {

"assigned_esi_level": 2,

"direction": "Emergency Department",

"confidence_level": 70,

"coded_explanation": {

"requires_immediate_lifesaving_intervention": 0,

"high_risk_situation": 0,

"confused_or_lethargic_or_disoriented": 0,

"severe_pain_or_distress": 1,

"vitals_in_the_danger_zone": 0,

"resources_needed_count": 3

}

}

},

{

"input": {

"age_years": null,

"time_of_presentation_iso8601": null,

"vital_parameters": {

"blood_pressure": "135/85 mmHg",

"heart_rate_bpm": 65,

"temperature_c": 36.2,

"blood_oxygen_percent": 98,

"breathing_rate_per_min": null

},

"additional_clinical_data": {

"airway_status": null,

"gcs": null,

"pain_scale_0_10": 6,

"blood_glucose_mg_dl": null,

"presence_of_vital_injury": "No"

},

"presenting_complaint": {

"cedis_code": "554",

"additional_notes": "Schmerzen in der Schulter rechts begonnen vor 4 Monaten beim Holz hacken, Ibu 600 und Novalgin eingenommen jetzt schlimmer"

}

},

"expected_output": {

"assigned_esi_level": 4,

"direction": "Emergency Department",

"confidence_level": 30,

"coded_explanation": {

"requires_immediate_lifesaving_intervention": 0,

"high_risk_situation": 0,

"confused_or_lethargic_or_disoriented": 0,

"severe_pain_or_distress": 0,

"vitals_in_the_danger_zone": 0,

"resources_needed_count": 1

}

}

},

{

"input": {

"age_years": null,

"time_of_presentation_iso8601": null,

"vital_parameters": {

"blood_pressure": "158/83 mmHg",

"heart_rate_bpm": 71,

"temperature_c": 36.2,

"blood_oxygen_percent": 100,

"breathing_rate_per_min": null

},

"additional_clinical_data": {

"airway_status": null,

"gcs": null,

"pain_scale_0_10": null,

"blood_glucose_mg_dl": null,

"presence_of_vital_injury": "No"

},

"presenting_complaint": {

"cedis_code": "861",

"additional_notes": "benötigt Asthaspray, kein Asthmaanfall, aber hat sein Spray beim Sport vergessen"

}

},

"expected_output": {

"assigned_esi_level": 5,

"direction": "Primary Care",

"confidence_level": 70,

"coded_explanation": {

"requires_immediate_lifesaving_intervention": 0,

"high_risk_situation": 0,

"confused_or_lethargic_or_disoriented": 0,

"severe_pain_or_distress": 0,

"vitals_in_the_danger_zone": 0,

"resources_needed_count": 0

}

}

}

]

}

**Prompt for c) ESI stepwise classification**

\## Model

gpt-5.1 reasoning high

\## Approach

This implementation uses a stepwise decision tree to classify emergency patients into ESI levels 1-5:

1. **Question A: Requires lifesaving intervention?**

- YES → ESI Level 1

- NO → Continue to Question B

2. **Question B: High-risk situation/confused/severe distress?**

- YES → ESI Level 2

- NO → Continue to Question C

3. **Question C: How many resource types needed?**

- Answers: None, One, or Many (2+)

- Continue to Question D

4. **Question D: High-risk vital signs?**

- Combined with Question C answer:

- None resources → ESI Level 5

- One resource + abnormal vitals → ESI Level 4

- One resource + normal vitals → ESI Level 5

- Many resources → ESI Level 3

Each question is posed to the LLM via a prompt that includes:

- All patient triage data (demographics, vitals, chief complaint, remarks)

- ESI guidelines specific to that decision step

- Structured answer options (YES/NO for A and B, NONE/ONE/MANY for C, YES/NO for D)

The model returns structured outputs (Pydantic models) with its answer and reasoning. The code then maps the answer to either:

- A specific ESI level (if terminal decision)

- The next question in the decision tree

\## Implementation

- Processes real triage data from CSV files

- Each patient goes through all applicable decision steps

- Outputs predictions with complete decision paths and reasoning

- Results saved as JSONL and CSV with ground truth for evaluation

- Visualization notebook shows decision tree with highlighted patient paths

\## Prompts Used

All prompts follow the same structure:

**System Message:**

```

You are an expert emergency medicine triage nurse for classifying patients based on the ESI (Emergency Severity Index) algorithm. Analyze the patient data and answer the triage question accurately. Base your decision on the clinical information provided. Use the provided guidelines of the ESI triage system to inform your answers.

```

**User Message:**

```

Patient Data:

{patient_data}

Question: {specific_question_with_guidelines}

```

\### Step A: Lifesaving Intervention

**Question:**

```

Does this patient require immediate lifesaving intervention?

Guidelines: Immediate life-saving intervention required: Airway or respiratory support, emergency medications, hemodynamic interventions such as fluid resuscitation or blood products. Clinical presentations requiring lifesaving interventions include the following: intubated, unresponsive, pulselessness, apneic, severe respiratory distress, profound hypotension or hypoglycemia. Unresponsiveness is defined as a patient who either:

1. Is nonverbal and not following commands (acutely)

OR

2. Requires noxious stimulus (P or U on AVPU scale)

```

\### Step B: High-Risk Situation

**Question:**

```

Is this a high-risk situation? (May become unstable, high deterioration risk, confused/lethargic/disoriented, or severe pain/distress)

Guidelines: High-risk situation: May become unstable, have high risk for deterioration, or exhibit newly altered mental status. Severe pain or distress is determined by patient report, corroborated with clinical observation.

```

\### Step C: Resource Needs

**Question:**

```

How many different types of resources does this patient need? (Count resource TYPES, not individual tests. Example: CBC + electrolytes + coagulation = ONE resource type [labs]. CBC + chest X-ray = TWO resource types [labs + imaging])

Guidelines: Resources: Count the number of different types of resources, not the individual tests or radiographs. (For example, complete blood count, electrolytes, and coagulant studies equal one resource because they are all laboratory tests, while complete blood count plus chest radiograph equals two resources because one is a laboratory test and one is imaging).

```

\### Step D: Vital Signs

**Question:**

```

Previous assessment found: {resource_count} resource(s) needed - {reasoning}

Does the patient have high-risk vital signs? Consider age-specific normal ranges and pediatric fever considerations.

Guidelines: High-risk vital signs: Reassess to determine whether the patient warrants a higher acuity level if a patient has one or more vital signs outside the normal parameters for the patient.

Oxygen saturation (SpO2) below 92% on room air is considered abnormal for all ages.

For pediatric patients, consider age-specific thresholds for heart rate (HR) and respiratory rate (RR). For patients > 18 years, HR > 100 bpm and RR > 20 breaths per minute are considered high-risk.

```
