## Supplementary material for "Performance evaluation and benchmarking across 16 large language model configurations on a large real-world emergency department triage data set": ANNEX: ANNEX_F_Sub-population_distribution.docx (from another version of this preprint: version 1, 2026-06-05)

**Annex F.a:** Nurse-assigned acuity distribution of encounters at the Central Point of Assessment.

| **ESI** | **Count (N = 2,000)** | **%** |
| --- | --- | --- |
| 1 | 1 | 0.05 |
| 2 | 213 | 10.65 |
| 3 | 742 | 37.1 |
| 4 | 765 | 38.25 |
| 5 | 279 | 13.95 |

**Annex F.b:** Nurse-assigned sectoral allocation at the Central Point of Assessment.

| **Sector** | **Count (N = 2,000)** | **%** |
| --- | --- | --- |
| ED | 1365 | 68.25 |
| UCP | 635 | 31.75 |

**Annex F.c:** Distribution of the 20 most common chief complaints at the Central Point of Assessment.

| **Chief Complaint** | **CEDIS code** | **Count (N = 2,000)** | **%** |
| --- | --- | --- | --- |
| Lower extremity pain | 555 | 166 | 8.3 |
| Abdominal Pain | 251 | 165 | 8.3 |
| Upper extremity pain | 554 | 141 | 7.05 |
| Backpain | 551 | 86 | 4.3 |
| Influenza-like illness | 065 | 69 | 3.45 |
| Upper respiratory infection symptoms | 066 | 62 | 3.1 |
| Upper extremity injury | 556 | 58 | 2.9 |
| Chest pain - cardiac features | 003 | 55 | 2.75 |
| Headache | 404 | 53 | 2.65 |
| Laceration/Puncture | 704 | 49 | 2.45 |
| Dental/gum problem | 101 | 48 | 2.4 |
| UTI complaints | 307 | 45 | 2.25 |
| Lower extremity injury | 557 | 43 | 2.15 |
| Sore throat | 103 | 36 | 1.8 |
| Neck swelling/pain | 104 | 36 | 1.8 |
| Shortness of breath | 006 | 35 | 1.75 |
| Localized swelling/redness | 709 | 35 | 1.75 |
| Chest pain - non-cardiac features | 004 | 32 | 1.6 |
| Fever | 852 | 30 | 1.5 |
| Nausea and/or vomiting | 251 | 29 | 1.45 |
| Other 105 complaints | n/a | 727 | 36 |

**Annex F.d:** Distribution of available clinical domains of the ABCDE framework(Thim et al. 2012) from the Triage Report Card (Annex B).

| **Clinical Domain** | **Count available (of N = 2,000)** | **% available** |
| --- | --- | --- |
| Airway | 1,104 | 55.2 |
| Breathing | 1,717 | 85.85 |
| Circulation | 1,758 | 87.9 |
| Disability | 224 | 11.2 |
| Exposure | 1,707 | 85.35 |
