## Supplementary figures and images for "Performance evaluation and benchmarking across 16 large language model configurations on a large real-world emergency department triage data set" (from another version of this preprint: version 1, 2026-06-05)

### ANNEX_G_Confidence_Distribution_Overview_2000_all_models.pdf

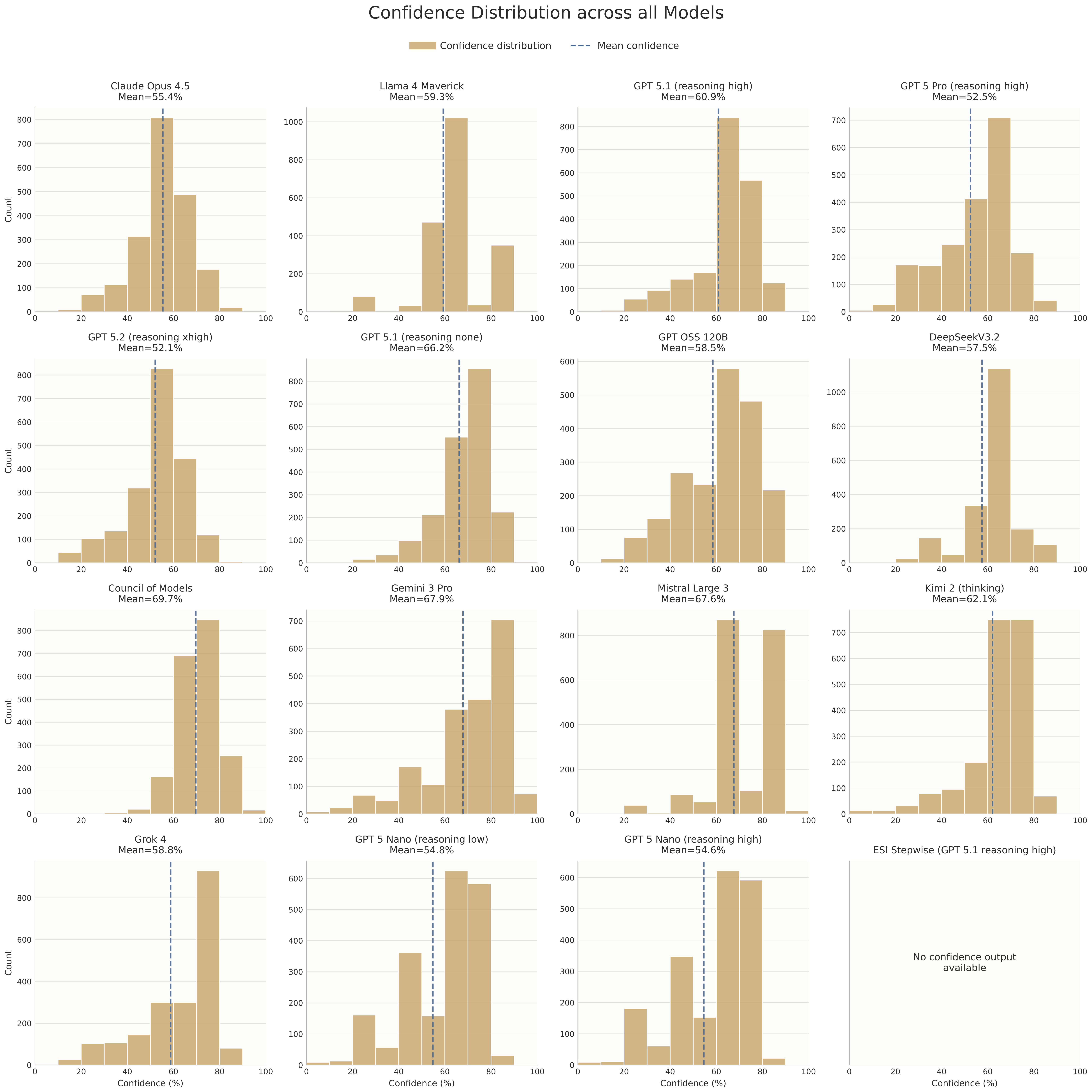

### ANNEX_H_Figure_Reliability_Overview_2000_all_models.pdf

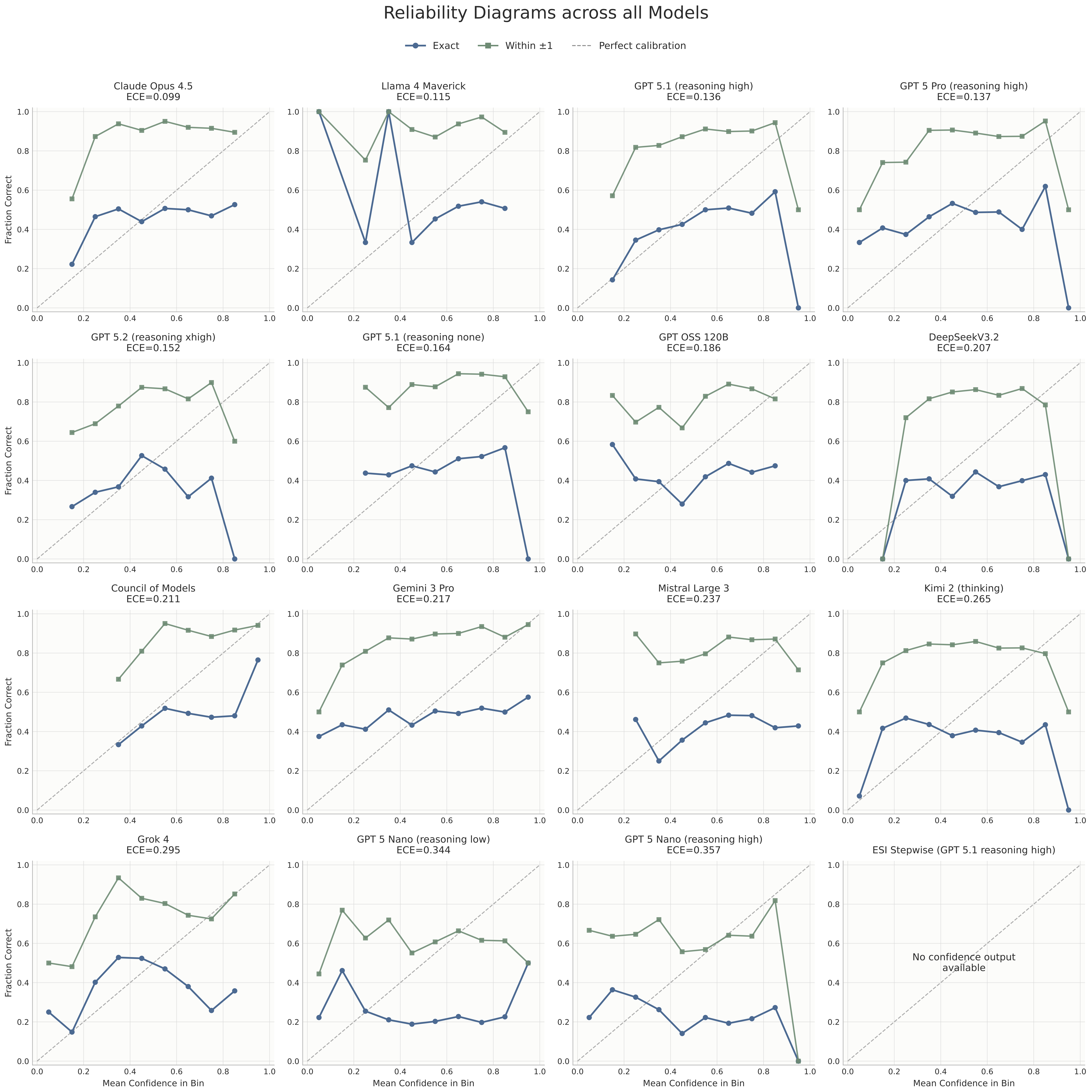

### ANNEX_I_Calibration_Companion_GPT_5_1_reasoning_high_16107.pdf

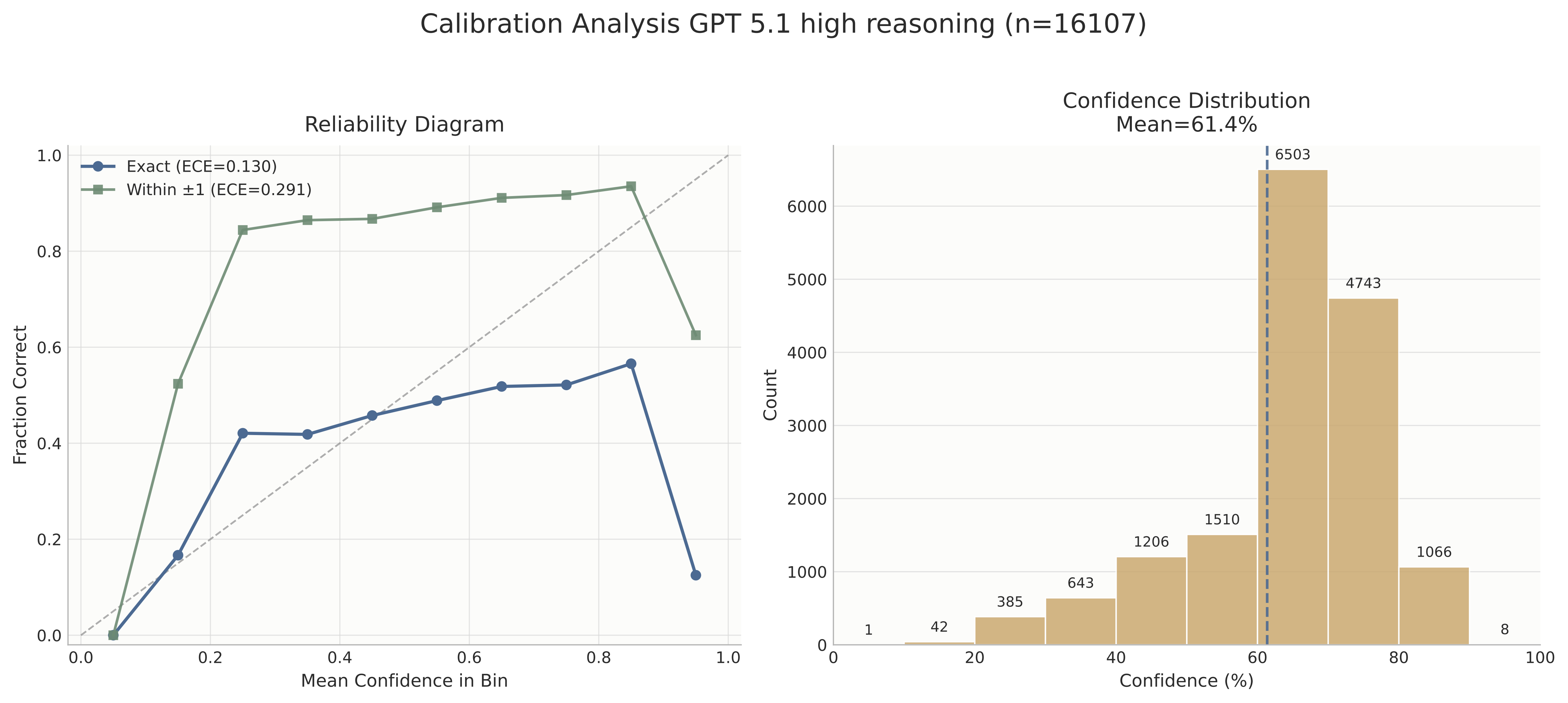
